# Environmental toxic metal exposure and conditional mortality survival among US adults: a prospective cohort study

**DOI:** 10.64898/2026.06.29.26356268

**Authors:** Xin Fang, Joel Schwartz

**Affiliations:** Department of Environmental Health, Harvard T.H. Chan School of Public Health, Boston, MA, USA

## Abstract

**Background:** Heavy metal exposure has been associated with mortality, but differences in early and median survival times remain unclear. We estimated associations of blood lead and mercury and urinary cadmium and arsenic at lower conditional survival-time quantiles.

**Methods:** We linked the 1999–2014 National Health and Nutrition Examination Survey to mortality records through Dec 31, 2019, including 24,338 adults aged 40 years or older. Weighted censored quantile regression estimated associations at the 10th, 25th, and 50th percentiles, adjusting for age and other confounders across five imputations. Mercury and arsenic analyses used 2003–2014 cycles; urinary-metal cohorts required observed estimated glomerular filtration rate ≥60 mL/min per 1.73 mZ.

**Findings:** Of 24,338 participants, 6,992 died. Doubling urinary cadmium was associated with 1.71–1.92, 1.50–1.67, and 1.35–1.53 years shorter conditional survival at the 10th, 25th, and 50th percentiles (n=7,061). Corresponding blood lead estimates were 0.63–0.71, 0.55–0.65, and 0.61–0.63 years shorter (n=24,021). Some median estimates were not estimable. Cadmium associations persisted after lead adjustment; lead associations did not persist after cadmium adjustment. Arsenic had no statistically significant association with mortality in this study. Mercury was associated with longer survival despite adjustment for fish or omega-3 intake, possibly reflecting residual confounding.

**Interpretation:** Low-to-moderate cadmium exposure was associated with shorter survival, with the largest absolute loss among adults who die earliest. These findings support continued efforts to reduce cadmium exposure, with particular benefit for adults most vulnerable to premature death.

**Funding:** None.

**Research in context:** *Evidence before this study:* We searched PubMed from database inception to Nov 10, 2026, without language restrictions, using metal names and biomarker terms combined with “mortality”, “survival”, “quantile regression”, “life expectancy”, and related terms. We prioritised prospective studies in adult populations and systematic reviews, supplemented by targeted searches for analyses of survival time. Previous studies have associated lead and cadmium exposure with higher all-cause and cardiovascular mortality. Evidence for mercury has varied across populations and exposure levels, and findings for low-level arsenic exposure in US adults have been less conclusive. Most cohort studies reported relative hazards, although earlier research examined metal-associated differences in longevity and used machine learning to investigate variation in lead-associated survival. Evidence remained limited on how associations with metal biomarkers differ between early and median conditional survival times in general populations.

*Added value of this study:* Among 24,338 US adults aged 40 years or older in NHANES 1999–2014, with mortality follow-up through 2019, we estimated associations of four metal biomarkers with the 10th, 25th, and 50th conditional percentiles of survival time after examination. Censored quantile regression provided estimands in years of life. In the urinary cadmium cohort of 7,061 participants with an estimated glomerular filtration rate of at least 60 mL/min per 1.73 mZ, a doubling of cadmium was associated with 1.71–1.92 years shorter to 10% mortality, compared with 1.35–1.53 years at the median. The cadmium associations persisted in NHANES III, although the largest estimate in that cohort was at the median. Lead and survival time associations were attenuated after adjustment for cadmium. Arsenic estimates were not significant. We observed positive association between blood mercury and survival time, which might reflect some residual confounding that we couldn’t control for.

*Implications of all the available evidence:* The evidence supports continued population-level efforts to reduce cadmium and lead exposure. With CQR, we were able to estimate the survival differences in years of life and for earlier deaths, providing valuable information for public health messages. Future studies using repeated exposure measurements and evaluations of exposure-reduction policies are needed.

## Introduction

Chronic exposure to the non-essential metals lead, cadmium, mercury, and inorganic arsenic has been associated with higher risks of cardiovascular disease, cancer, and kidney disease.^1–3^ One study attributed about one-third of the decline in US cardiovascular mortality between 1988– 1994 and 1999–2004 to reductions in lead and cadmium exposure.^4^ Although exposure to lead and cadmium has fallen in the USA, both metals remain important determinants of population health and mortality at current concentrations.^5^

The US National Health and Nutrition Examination Survey (NHANES) provides evidence on mortality at these exposure levels. Comparing the 80th with the 20th percentile of urinary cadmium in NHANES 1999–2004, Téllez-Plaza and colleagues reported hazard ratios of 1.52 (95% CI 1.00–2.29) for all-cause mortality and 1.74 (1.07–2.83) for cardiovascular mortality.^6^ In NHANES III, Lanphear and colleagues estimated that 18.0% (95% CI 10.9–26.1) of all deaths and 28.7% (15.5–39.5) of cardiovascular deaths were attributable to lead exposure.^7^ Inorganic arsenic, encountered mainly through water and food, has been associated to cardiovascular disease risk,^8^ whereas evidence on mercury and mortality remains mixed.^1,9,10^

Whether exposure to these metals shortens the lives of people already vulnerable to premature death is unknown. Previous studies have generally reported hazard ratios, which compare death rates among people still alive at each point and are assumed to be constant during follow-up. As deaths accumulate, however, the remaining participants are likely to be healthier or genetically predisposed to live longer despite their exposure.^11,12^ Among adults of the same age at enrolment, it is therefore unclear how much sooner the earliest deaths occur at higher metal exposures, and whether this difference is larger or smaller than at the median survival time. Censored quantile regression can help answer this public health question by estimating differences in survival time across the survival distribution.^13,14^ For adults of the same baseline age, the 10th percentile is the time after examination by which 10% are expected to have died in years.

We linked eight cycles of NHANES (1999–2014) to the National Death Index through Dec 31, 2019. Among US adults aged 40 years or older, we estimated how a doubling of whole-blood lead, whole-blood total mercury, urinary cadmium, and the sum of urinary inorganic and methylated arsenic species was associated with the times by which 10%, 25%, and 50% of participants had died. We hypothesized that higher metal concentrations would be associated with a larger reduction in time to death to 10% mortality than in time to death to 50% mortality. For lead and cadmium, we also examined dose–response curves.

## Methods

### Study design and participants

We conducted a prospective cohort study using eight cycles (1999–2014) of the continuous National Health and Nutrition Examination Survey (NHANES), linked to the National Death Index (NDI) with follow-up through Dec 31, 2019. NHANES uses a complex, stratified, multistage probability sample of the civilian, non-institutionalised US population, with oversampling of older adults and of non-Hispanic Black and Hispanic individuals.^15^ The National Center for Health Statistics (NCHS) Research Ethics Review Board approved the NHANES protocols, and all participants provided written informed consent. This secondary analysis of de-identified, publicly available data was determined to be non-human-subjects research and exempt from institutional review. This study is reported according to the STROBE guideline.

Eligible participants were non-pregnant adults aged 40 years or older at the Mobile Examination Center (MEC) visit who were eligible for mortality follow-up and had at least one of the four metal biomarkers measured; 24,338 of 82,091 NHANES 1999–2014 participants (6,992 deaths) met these criteria (figure 1). The metal-specific primary cohorts comprised 24,021 participants (6,885 deaths) for whole-blood lead (1999–2014), 18,258 (4,358 deaths) for whole-blood total mercury (2003–2014), 7,061 (1,510 deaths) for urinary cadmium (1999–2014), and 5,552 (960 deaths) for urinary arsenic (2003–2014). The urinary cohorts were smaller because urinary metals were measured in a one-third subsample and because we restricted them to participants with observed estimated glomerular filtration rate (eGFR) of 60 mL/min per 1.73 mZ or higher, excluding 973 and 779 participants with eGFR below 60 and 432 and 338 with missing eGFR, respectively.

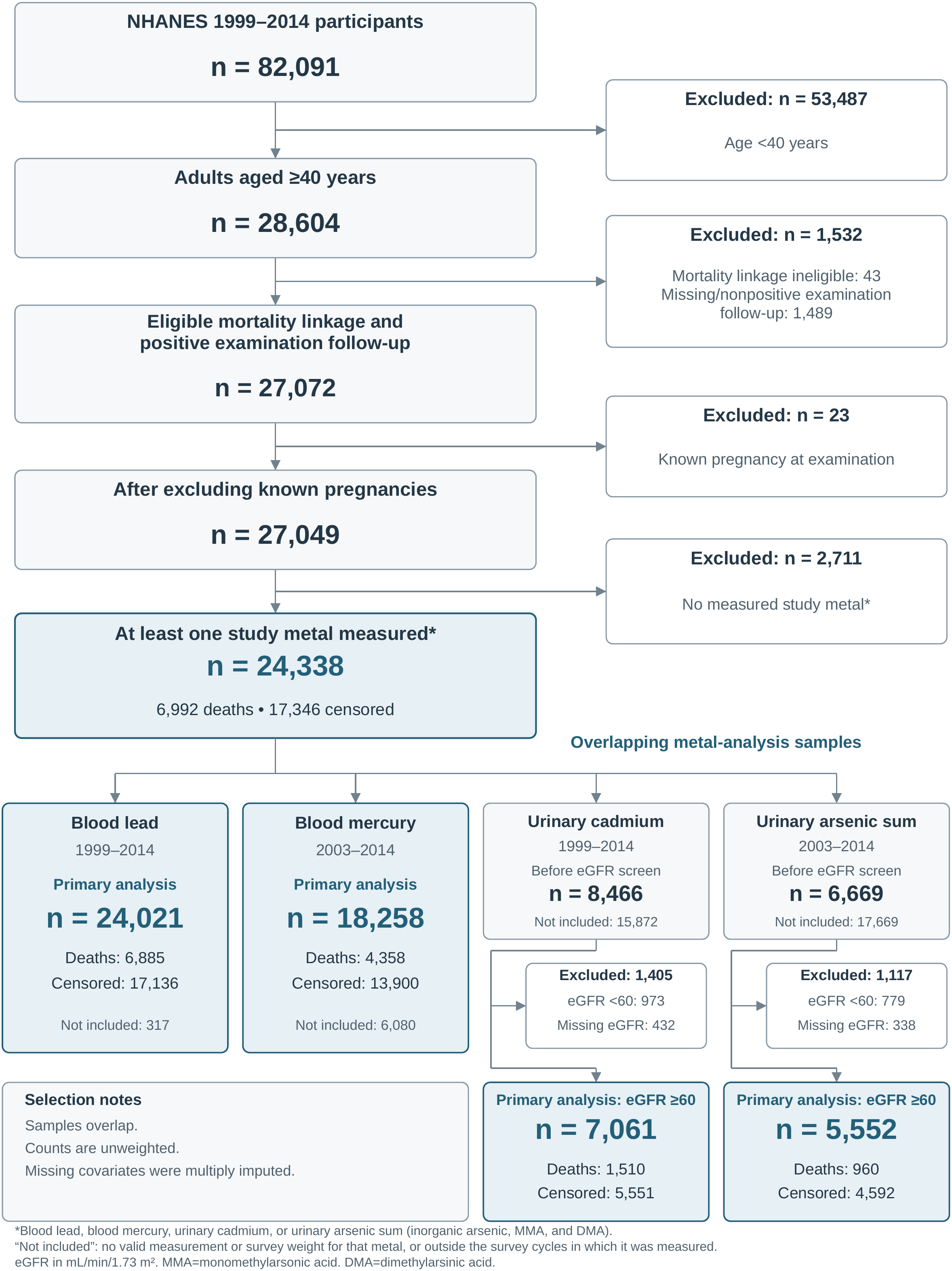

### Exposure assessment

The four primary exposures were whole-blood lead (µg/dL), whole-blood total mercury (µg/L), urinary cadmium (µg/L), and the sum of inorganic, monomethylated, and dimethylated urinary arsenic species (µg/L). Blood lead was measured by graphite-furnace atomic absorption spectrometry in 1999–2002 and inductively coupled plasma mass spectrometry (ICP-MS) thereafter. Total blood mercury and urinary cadmium were measured by ICP-MS; urinary arsenic species were separated by high-performance liquid chromatography coupled to ICP-MS with a dynamic reaction cell (appendix p 26). Values below detection limits were kept as LOD/√2.

To account for urine dilution, we applied the covariate-adjusted creatinine standardisation of O’Brien and colleagues^16^ to urinary cadmium and arsenic instead of conventional division by creatinine, which can induce collider bias when creatinine is influenced by covariates that also affect survival time. Each metal was then log₂-transformed so that regression coefficients are interpretable as the change in years of survival time per doubling of metal concentration.

### Outcome

The primary outcome was all-cause mortality, ascertained from the NDI-linked mortality file. Survival time was calculated from the MEC examination to death or Dec 31, 2019, and expressed in years.

### Covariates and causal structure

A directed acyclic graph informed confounders selection (appendix p 4). All models adjusted for age (linear and quadratic), sex, self-reported race and ethnicity, education, marital status, income-to-poverty ratio, pack-years of smoking, alcohol use (lifetime abstainer, former, light, moderate, or heavy), cycle-specific physical-activity rank groups, NHANES cycle, body-mass index (BMI), and total cholesterol. Because reduced kidney function raises urinary metal concentrations and shortens survival, we restricted the urinary-metal cohorts to participants with observed eGFR of 60 mL/min per 1.73 mZ or higher at the design stage and adjusted for continuous eGFR, calculated with the 2021 race-free CKD-EPI creatinine equation,^17^ at the analysis stage.

Fatty fish intake is a major source of mercury exposure^18^ and provides long-chain omega-3 fatty acids with cardiovascular and anti-inflammatory benefits.^19,20^ Mercury models therefore adjusted for fish intake, prioritising a 30-day species screener (Tier 1; n=14,757), followed by reliable 24-h recalls (Tier 2; n=2,505), within the 2003–2014 cohort (appendix p 29). Missing intake (996 participants; 4.9%, survey weighted) was multiply imputed.

### Statistical analysis

We fitted censored quantile regression (CQR) with the Peng–Huang estimator^14^ in the R package quantreg. Coefficients represent differences in conditional survival-time quantiles, in years, per doubling of the metal biomarkers, holding covariates fixed. For example, a coefficient of −1 represents 1-year shorter survival until death at that quantile. Because the Peng–Huang estimator identifies quantiles only up to an upper limit set by censoring, and our question concerns the earliest deaths, τ = 0.10 and τ = 0.25 were primary and the median was secondary. Quantiles that are inestimable in each imputation are reported as not estimable (NE).

Point estimates used metal-specific sampling weights, combined across cycles following NCHS guidance^15^ and normalised to the cohort mean (appendix p 2). Missing covariates were multiply imputed by chained equations (five imputations, 20 iterations).^21^ Primary confidence intervals used 400 delete-d jackknife resamples per imputation. Analyses were done in R (version 4.6.1) with the packages quantreg (version 6.1), survival, survey, and mice.^21^

### Dose–response

To explore exposure–response shapes for lead and cadmium, the metals with the largest associations, we fitted four-knot restricted cubic splines for log₂-transformed exposure, with knots at the unweighted 5th, 35th, 65th, and 95th percentiles.^22^ Models retained the corresponding primary covariates and weights, including continuous eGFR for cadmium.

### Sensitivity analyses

Four sensitivity analyses were prespecified: using an alternative Portnoy’s recursively reweighted estimator;^13^ restricting to participants aged 55 years or older; mutually adjusting blood lead and urinary cadmium among participants with both measurements; and repeating the lead and cadmium analyses in an independent cohort, NHANES III (1988–1994; follow-up through Dec 31, 2019). Sensitivity analyses of the urinary metals were restricted to participants with eGFR of 60 mL/min per 1.73 mZ or higher, as in the primary analysis. In a post-hoc analysis, we replaced the fish covariate in the mercury model with total dietary long-chain omega-3 fatty acid (EPA plus DHA) intake.

### Effect modification by nutrient biomarkers

We performed a literature review (appendix pp 2, 10) on mechanistically plausible nutrient biomarkers that could potentially modify the association between heavy metals and mortality. Informed by the review, we conducted exploratory analyses on whether serum vitamin C, serum α-carotene, and red blood cell (RBC) folate modified the metal–mortality associations on the additive scale by adding a metal × biomarker product term to the primary models. P values for the 12 primary interaction tests at τ = 0.10 were corrected with the Benjamini–Hochberg procedure.

### Cancer and cardiovascular mortality

In secondary analyses, we examined whether heavy metal was associated with cancer and cardiovascular mortality for lower tail of the survival distribution. Interpretation requires competing-event censoring to be independent of survival time until death conditional on covariates. Fewer quantiles were estimable because each specific cause contributed fewer deaths. Outcome codes and details are given in the appendix (pp 3, 16-18).

## Results

### Cohort characteristics

The analytic population comprised 24,338 adults, of whom 6,992 died by Dec 31, 2019 (figure 1). The four metal cohorts had similar characteristics (table 1). In the blood lead cohort (n = 24,021; 6,885 deaths), the survey-weighted median age at examination was 55 years (IQR 47– 66), 52.9% were female, and 74.9% were non-Hispanic White. Median follow-up ranged from 9.8 years (IQR 6.8–13.0) for mercury to 11.2 years (7.8–15.2) for cadmium. Median baseline concentrations were 1.57 µg/dL (IQR 1.03–2.33) for whole-blood lead, 1.00 µg/L (0.52–1.97) for whole-blood total mercury, 0.29 µg/L (0.17–0.49) for creatinine-standardised urinary cadmium, and 5.89 µg/L (4.02–8.99) for urinary arsenic, consistent with the low-to-moderate exposures reported for US adults over the past two decades.^23^ Covariate missingness was at most 7.2% (appendix p 8).

**Table 1.**
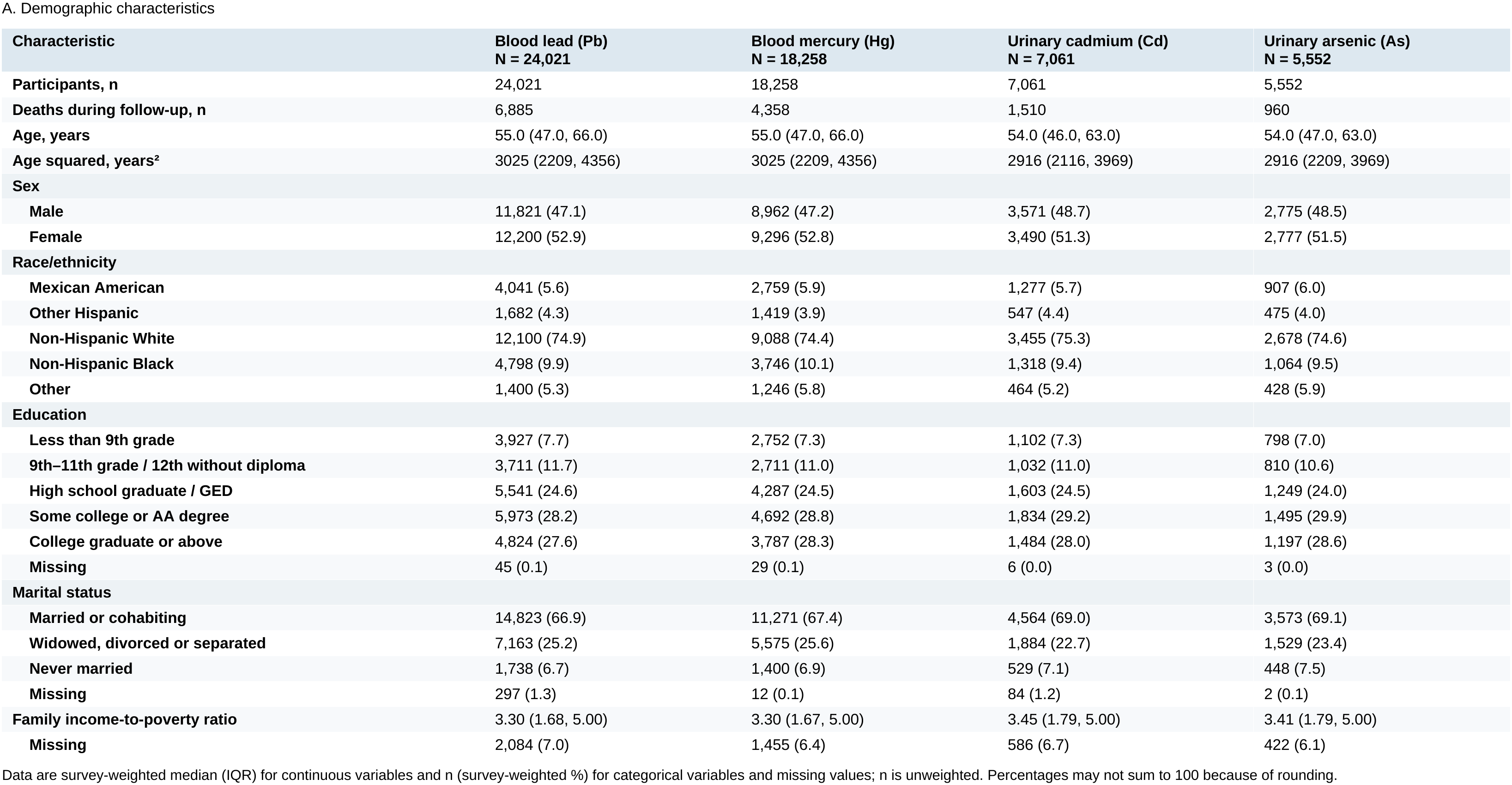
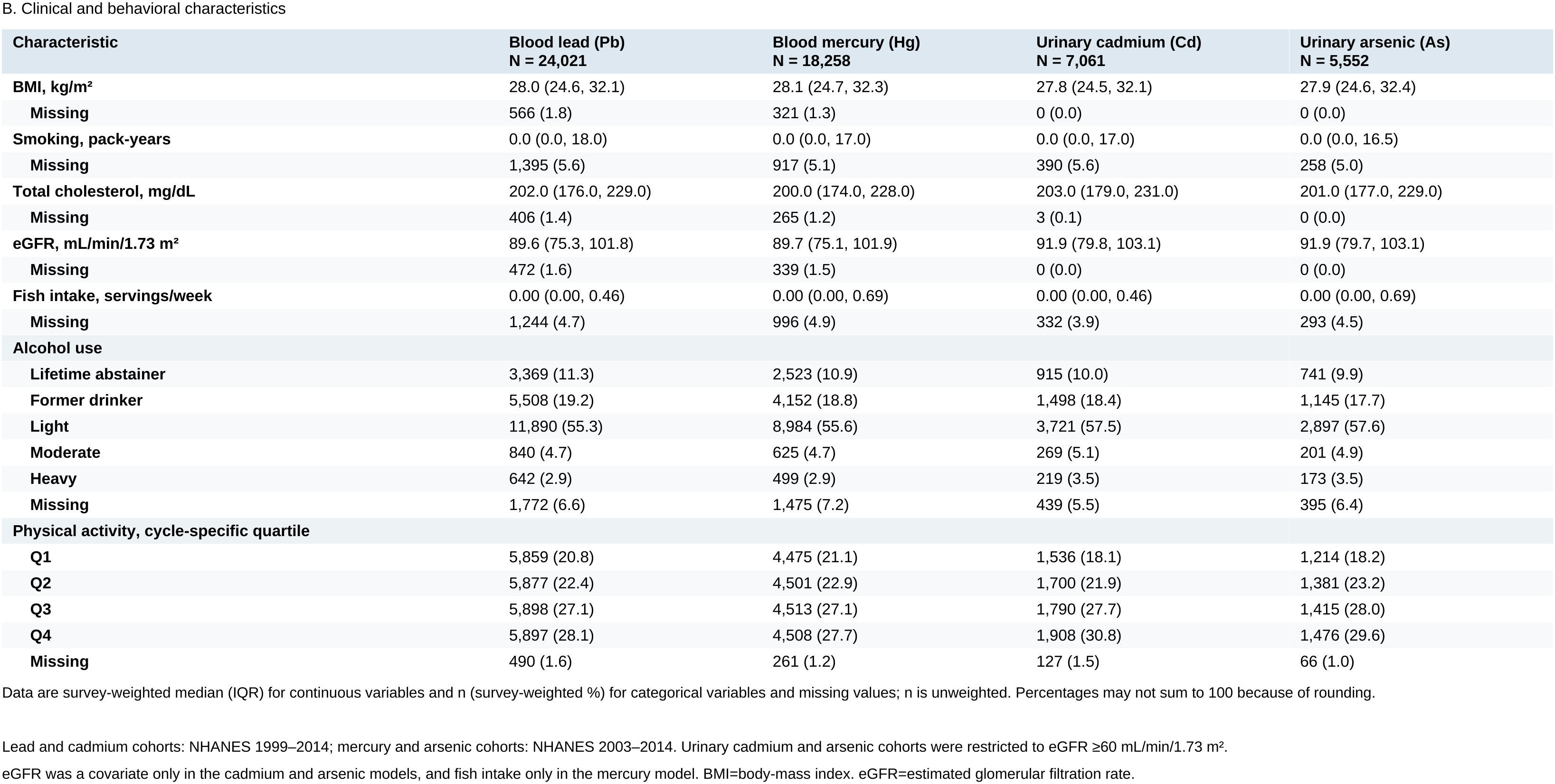
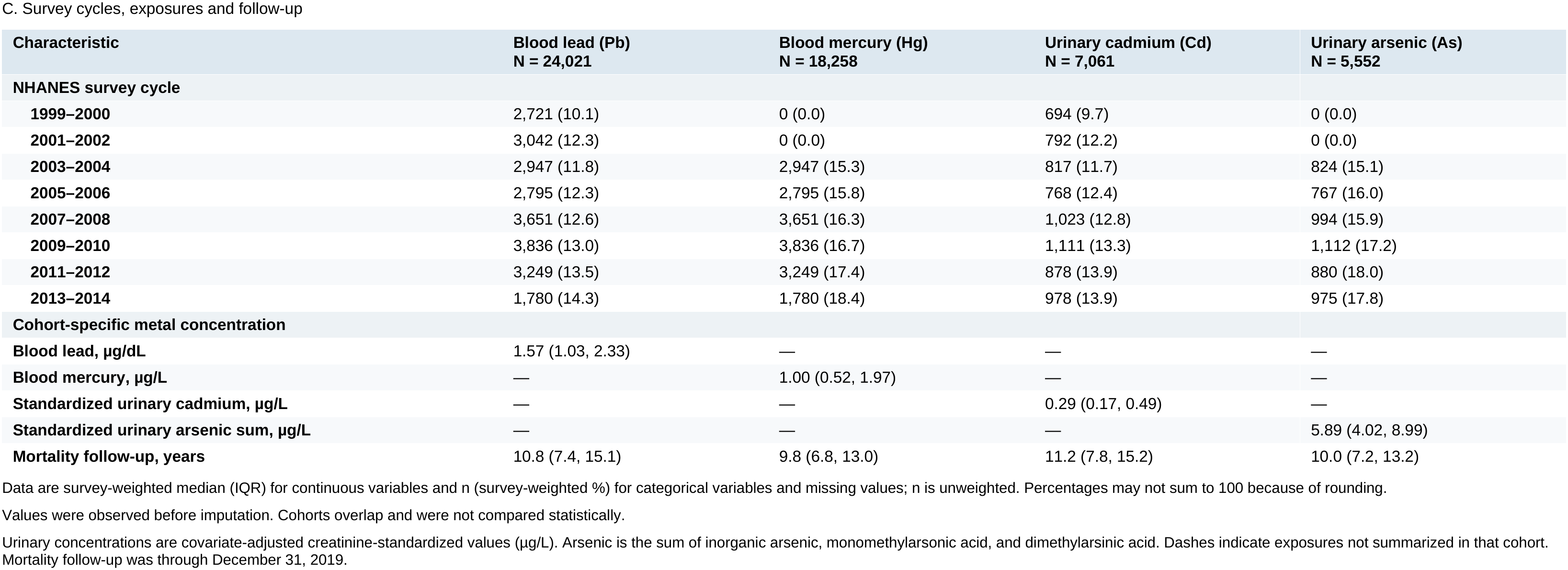
Baseline characteristics of the four primary metal cohorts.

| **Characteristic** | **Blood lead (Pb)**<br>**N = 24,021** | **Blood mercury (Hg)**<br>**N = 18,258** | **Urinary cadmium (Cd)**<br>**N = 7,061** | **Urinary arsenic (As)**<br>**N = 5,552** |
|---|---|---|---|---|
| **Participants, n** | 24,021 | 18,258 | 7,061 | 5,552 |
| **Deaths during follow-up, n** | 6,885 | 4,358 | 1,510 | 960 |
| **Age, years** | 55.0 (47.0, 66.0) | 55.0 (47.0, 66.0) | 54.0 (46.0, 63.0) | 54.0 (47.0, 63.0) |
| **Age squared, years<sup>2</sup>** | 3025 (2209, 4356) | 3025 (2209, 4356) | 2916 (2116, 3969) | 2916 (2209, 3969) |
| **Sex** |  |  |  |  |
| **Male** | 11,821 (47.1) | 8,962 (47.2) | 3,571 (48.7) | 2,775 (48.5) |
| **Female** | 12,200 (52.9) | 9,296 (52.8) | 3,490 (51.3) | 2,777 (51.5) |
| **Race/ethnicity** |  |  |  |  |
| **Mexican American** | 4,041 (5.6) | 2,759 (5.9) | 1,277 (5.7) | 907 (6.0) |
| **Other Hispanic** | 1,682 (4.3) | 1,419 (3.9) | 547 (4.4) | 475 (4.0) |
| **Non-Hispanic White** | 12,100 (74.9) | 9,088 (74.4) | 3,455 (75.3) | 2,678 (74.6) |
| **Non-Hispanic Black** | 4,798 (9.9) | 3,746 (10.1) | 1,318 (9.4) | 1,064 (9.5) |
| **Other** | 1,400 (5.3) | 1,246 (5.8) | 464 (5.2) | 428 (5.9) |
| **Education** |  |  |  |  |
| **Less than 9th grade** | 3,927 (7.7) | 2,752 (7.3) | 1,102 (7.3) | 798 (7.0) |
| **9th–11th grade / 12th without diploma** | 3,711 (11.7) | 2,711 (11.0) | 1,032 (11.0) | 810 (10.6) |
| **High school graduate / GED** | 5,541 (24.6) | 4,287 (24.5) | 1,603 (24.5) | 1,249 (24.0) |
| **Some college or AA degree** | 5,973 (28.2) | 4,692 (28.8) | 1,834 (29.2) | 1,495 (29.9) |
| **College graduate or above** | 4,824 (27.6) | 3,787 (28.3) | 1,484 (28.0) | 1,197 (28.6) |
| **Missing** | 45 (0.1) | 29 (0.1) | 6 (0.0) | 3 (0.0) |
| **Marital status** |  |  |  |  |
| **Married or cohabiting** | 14,823 (66.9) | 11,271 (67.4) | 4,564 (69.0) | 3,573 (69.1) |
| **Widowed, divorced or separated** | 7,163 (25.2) | 5,575 (25.6) | 1,884 (22.7) | 1,529 (23.4) |
| **Never married** | 1,738 (6.7) | 1,400 (6.9) | 529 (7.1) | 448 (7.5) |
| **Missing** | 297 (1.3) | 12 (0.1) | 84 (1.2) | 2 (0.1) |
| **Family income-to-poverty ratio** | 3.30 (1.68, 5.00) | 3.30 (1.67, 5.00) | 3.45 (1.79, 5.00) | 3.41 (1.79, 5.00) |
| **Missing** | 2,084 (7.0) | 1,455 (6.4) | 586 (6.7) | 422 (6.1) |
| **Characteristic** | **Blood lead (Pb)**<br>**N = 24,021** | **Blood mercury (Hg)**<br>**N = 18,258** | **Urinary cadmium (Cd)**<br>**N = 7,061** | **Urinary arsenic (As)**<br>**N = 5,552** |
| **BMI, kg/m<sup>2</sup>** | 28.0 (24.6, 32.1) | 28.1 (24.7, 32.3) | 27.8 (24.5, 32.1) | 27.9 (24.6, 32.4) |
| **Missing** | 566 (1.8) | 321 (1.3) | 0 (0.0) | 0 (0.0) |
| **Smoking, pack-years** | 0.0 (0.0, 18.0) | 0.0 (0.0, 17.0) | 0.0 (0.0, 17.0) | 0.0 (0.0, 16.5) |
| **Missing** | 1,395 (5.6) | 917 (5.1) | 390 (5.6) | 258 (5.0) |
| **Total cholesterol, mg/dL** | 202.0 (176.0, 229.0) | 200.0 (174.0, 228.0) | 203.0 (179.0, 231.0) | 201.0 (177.0, 229.0) |
| **Missing** | 406 (1.4) | 265 (1.2) | 3 (0.1) | 0 (0.0) |
| **eGFR, mL/min/1.73 m<sup>2</sup>** | 89.6 (75.3, 101.8) | 89.7 (75.1, 101.9) | 91.9 (79.8, 103.1) | 91.9 (79.7, 103.1) |
| **Missing** | 472 (1.6) | 339 (1.5) | 0 (0.0) | 0 (0.0) |
| **Fish intake, servings/week** | 0.00 (0.00, 0.46) | 0.00 (0.00, 0.69) | 0.00 (0.00, 0.46) | 0.00 (0.00, 0.69) |
| **Missing** | 1,244 (4.7) | 996 (4.9) | 332 (3.9) | 293 (4.5) |
| **Alcohol use** |  |  |  |  |
| **Lifetime abstainer** | 3,369 (11.3) | 2,523 (10.9) | 915 (10.0) | 741 (9.9) |
| **Former drinker** | 5,508 (19.2) | 4,152 (18.8) | 1,498 (18.4) | 1,145 (17.7) |
| **Light** | 11,890 (55.3) | 8,984 (55.6) | 3,721 (57.5) | 2,897 (57.6) |
| **Moderate** | 840 (4.7) | 625 (4.7) | 269 (5.1) | 201 (4.9) |
| **Heavy** | 642 (2.9) | 499 (2.9) | 219 (3.5) | 173 (3.5) |
| **Missing** | 1,772 (6.6) | 1,475 (7.2) | 439 (5.5) | 395 (6.4) |
| **Physical activity, cycle-specific quartile** |  |  |  |  |
| **Q1** | 5,859 (20.8) | 4,475 (21.1) | 1,536 (18.1) | 1,214 (18.2) |
| **Q2** | 5,877 (22.4) | 4,501 (22.9) | 1,700 (21.9) | 1,381 (23.2) |
| **Q3** | 5,898 (27.1) | 4,513 (27.1) | 1,790 (27.7) | 1,415 (28.0) |
| **Q4** | 5,897 (28.1) | 4,508 (27.7) | 1,908 (30.8) | 1,476 (29.6) |
| **Missing** | 490 (1.6) | 261 (1.2) | 127 (1.5) | 66 (1.0) |
| **Characteristic** | **Blood lead (Pb)**<br>**N = 24,021** | **Blood mercury (Hg)**<br>**N = 18,258** | **Urinary cadmium (Cd)**<br>**N = 7,061** | **Urinary arsenic (As)**<br>**N = 5,552** |
| **NHANES survey cycle** |  |  |  |  |
| **1999–2000** | 2,721 (10.1) | 0 (0.0) | 694 (9.7) | 0 (0.0) |
| **2001–2002** | 3,042 (12.3) | 0 (0.0) | 792 (12.2) | 0 (0.0) |
| **2003–2004** | 2,947 (11.8) | 2,947 (15.3) | 817 (11.7) | 824 (15.1) |
| **2005–2006** | 2,795 (12.3) | 2,795 (15.8) | 768 (12.4) | 767 (16.0) |
| **2007–2008** | 3,651 (12.6) | 3,651 (16.3) | 1,023 (12.8) | 994 (15.9) |
| **2009–2010** | 3,836 (13.0) | 3,836 (16.7) | 1,111 (13.3) | 1,112 (17.2) |
| **2011–2012** | 3,249 (13.5) | 3,249 (17.4) | 878 (13.9) | 880 (18.0) |
| **2013–2014** | 1,780 (14.3) | 1,780 (18.4) | 978 (13.9) | 975 (17.8) |
| **Cohort-specific metal concentration** |  |  |  |  |
| **Blood lead, µg/dL** | 1.57 (1.03, 2.33) | — | — | — |
| **Blood mercury, µg/L** | — | 1.00 (0.52, 1.97) | — | — |
| **Standardized urinary cadmium, µg/L** | — | — | 0.29 (0.17, 0.49) | — |
| **Standardized urinary arsenic sum, µg/L** | — | — | — | 5.89 (4.02, 8.99) |
| **Mortality follow-up, years** | 10.8 (7.4, 15.1) | 9.8 (6.8, 13.0) | 11.2 (7.8, 15.2) | 10.0 (7.2, 13.2) |

Data are survey-weighted median (IQR) for continuous variables and n (survey-weighted %) for categorical variables and missing values; n is unweighted. Percentages may not sum to 100 because of rounding.
Data are survey-weighted median (IQR) for continuous variables and n (survey-weighted %) for categorical variables and missing values; n is unweighted. Percentages may not sum to 100 because of rounding.
Lead and cadmium cohorts: NHANES 1999–2014; mercury and arsenic cohorts: NHANES 2003–2014. Urinary cadmium and arsenic cohorts were restricted to eGFR ≥60 mL/min/1.73 m<sup>2</sup>.
eGFR was a covariate only in the cadmium and arsenic models, and fish intake only in the mercury model. BMI=body-mass index. eGFR=estimated glomerular filtration rate.
Data are survey-weighted median (IQR) for continuous variables and n (survey-weighted %) for categorical variables and missing values; n is unweighted. Percentages may not sum to 100 because of rounding.
Values were observed before imputation. Cohorts overlap and were not compared statistically.
Urinary concentrations are covariate-adjusted creatinine-standardized values (µg/L). Arsenic is the sum of inorganic arsenic, monomethylarsonic acid, and dimethylarsinic acid. Dashes indicate exposures not summarized in that cohort. Mortality follow-up was through December 31, 2019.

### Primary analysis

A doubling of urinary cadmium (eg, from the median of 0.29 µg/L to 0.58 µg/L) was associated with 1.71–1.92 fewer years of survival time at the 10th percentile, 1.50–1.67 fewer years at the 25th percentile, and 1.35–1.53 fewer years at the median across the five imputations (n = 7,061; 1,510 deaths; all p ≤ 0.0017; table 2). The absolute reduction was largest at τ = 0.10, the lower tail of the survival distribution.

**Table 2.** Association of metal concentrations with conditional quantiles of survival time, by imputation.

| Metal | 10th percentile | 25th percentile | 50th percentile |
|---|---|---|---|
| **Blood lead (Pb)**<br>**N=24,021; deaths=6,885** | I1: −0.671 (−0.906, −0.411); P<0.0001<br>I2: −0.685 (−0.923, −0.437); P<0.0001<br>I3: −0.679 (−0.995, −0.354); P<0.0001<br>I4: −0.706 (−0.974, −0.461); P<0.0001<br>I5: −0.625 (−0.890, −0.310); P<0.0001 | I1: −0.555 (−0.916, −0.140); P=0.0051<br>I2: −0.607 (−0.813, −0.380); P<0.0001<br>I3: −0.587 (−0.817, −0.365); P<0.0001<br>I4: −0.650 (−0.895, −0.388); P<0.0001<br>I5: −0.612 (−0.887, −0.268); P=0.0001 | I1: NE<br>I2: −0.613 (−0.815, −0.383); P<0.0001<br>I3: −0.626 (−0.879, −0.281); P<0.0001<br>I4: NE<br>I5: −0.620 (−0.913, −0.319); P<0.0001 |
| **Blood mercury (Hg)**<br>**N=18,258; deaths=4,358** | I1: 0.328 (0.168, 0.487); P<0.0001<br>I2: 0.286 (0.172, 0.420); P<0.0001<br>I3: 0.327 (0.160, 0.483); P<0.0001<br>I4: 0.287 (0.098, 0.456); P=0.0016<br>I5: 0.327 (0.165, 0.524); P=0.0003 | I1: 0.477 (0.320, 0.600); P<0.0001<br>I2: NE<br>I3: 0.511 (0.312, 0.704); P<0.0001<br>I4: 0.502 (0.307, 0.689); P<0.0001<br>I5: 0.529 (0.315, 0.813); P<0.0001 | I1: 0.572 (0.336, 0.797); P<0.0001<br>I2: NE<br>I3: NE<br>I4: 0.579 (0.378, 0.776); P<0.0001<br>I5: 0.538 (0.275, 0.714); P<0.0001 |
| **Urinary cadmium (Cd)**<br>**N=7,061; deaths=1,510** | I1: −1.763 (−2.514, −1.008); P<0.0001<br>I2: −1.708 (−2.535, −0.962); P<0.0001<br>I3: −1.723 (−2.480, −1.032); P<0.0001<br>I4: −1.741 (−2.365, −1.131); P<0.0001<br>I5: −1.919 (−2.534, −1.140); P<0.0001 | I1: −1.638 (−2.117, −1.117); P<0.0001<br>I2: −1.571 (−2.134, −0.908); P<0.0001<br>I3: −1.501 (−1.963, −0.865); P<0.0001<br>I4: −1.632 (−2.209, −1.070); P<0.0001<br>I5: −1.665 (−2.090, −1.025); P<0.0001 | I1: −1.347 (−1.938, −0.755); P<0.0001<br>I2: −1.521 (−2.135, −0.890); P<0.0001<br>I3: −1.508 (−2.103, −0.894); P<0.0001<br>I4: −1.535 (−2.234, −0.945); P<0.0001<br>I5: −1.395 (−2.375, −0.628); P=0.0017 |
| **Urinary arsenic (As)**<br>**N=5,552; deaths=960** | I1: 0.177 (−0.379, 0.772); P=0.5468<br>I2: 0.321 (−0.430, 1.089); P=0.4070<br>I3: 0.058 (−0.655, 1.049); P=0.8941<br>I4: 0.075 (−0.624, 0.792); P=0.8364<br>I5: 0.035 (−0.540, 0.603); P=0.9035 | I1: 0.327 (−0.775, 1.151); P=0.5058<br>I2: 0.416 (−0.293, 1.220); P=0.2818<br>I3: 0.541 (−0.183, 1.264); P=0.1426<br>I4: 0.536 (−0.205, 1.349); P=0.1763<br>I5: 0.466 (−0.644, 1.369); P=0.3642 | I1: NE<br>I2: 0.289 (−0.780, 1.368); P=0.5979<br>I3: 0.300 (−0.341, 1.056); P=0.3994<br>I4: NE<br>I5: NE |

Values are the difference in the conditional quantile of survival time (years) per doubling of metal concentration (95% CI) and P value, shown separately for each of five imputations (I1–I5). Column percentiles refer to the conditional distribution of survival time; negative values indicate shorter survival time. NE=not estimable.
Urinary cadmium and arsenic cohorts were restricted to eGFR ≥60 mL/min/1.73 m². 95% CIs were estimated from 400 delete-d jackknife resamples and are not pooled across imputations or adjusted for the survey design.
Models were adjusted for age, age squared, sex, race and ethnicity, education, marital status, income-to-poverty ratio, BMI, smoking pack-years, alcohol use, physical activity, survey cycle, and total cholesterol; cadmium and arsenic models were additionally adjusted for eGFR, and the mercury model for fish intake. BMI=body-mass index. eGFR=estimated glomerular filtration rate.

A doubling of whole-blood lead (eg, from 1.57 µg/dL to 3.14 µg/dL) was associated with 0.63–0.71 fewer years at the 10th percentile, 0.55–0.65 at the 25th percentile, and 0.61–0.63 at the median (n = 24,021; 6,885 deaths; all p ≤ 0.0051), with the largest reduction at τ = 0.10; the median was not estimable in two imputations (table 2). This marginal association did not persist after mutual adjustment for cadmium.

Urinary arsenic was not associated with survival time at any quantile (β 0.04 to 0.32 years at τ = 0.10 and 0.33 to 0.54 years at τ = 0.25; all 95% CIs included zero; p ≥ 0.14; n = 5,552; 960 deaths); the median was estimable in two imputations only (table 2).

Whole-blood total mercury was positively associated with survival time at all three quantiles: a doubling (eg, from 1.00 µg/L to 2.00 µg/L) was associated with 0.29–0.33 more years at τ = 0.10, 0.48–0.53 at τ = 0.25 (four imputations), and 0.54–0.58 at τ = 0.50 (three imputations; n = 18,258; 4,358 deaths; table 2).

### Dose–response analysis

For urinary cadmium, predicted survival time declined across the exposure range at all three quantiles (figure 2). At τ = 0.10, predicted survival time was 5.08–5.90 years shorter at the 95th than at the 5th percentile of cadmium concentration across the four imputations with an available curve (appendix p 3).

**Figure 2.**
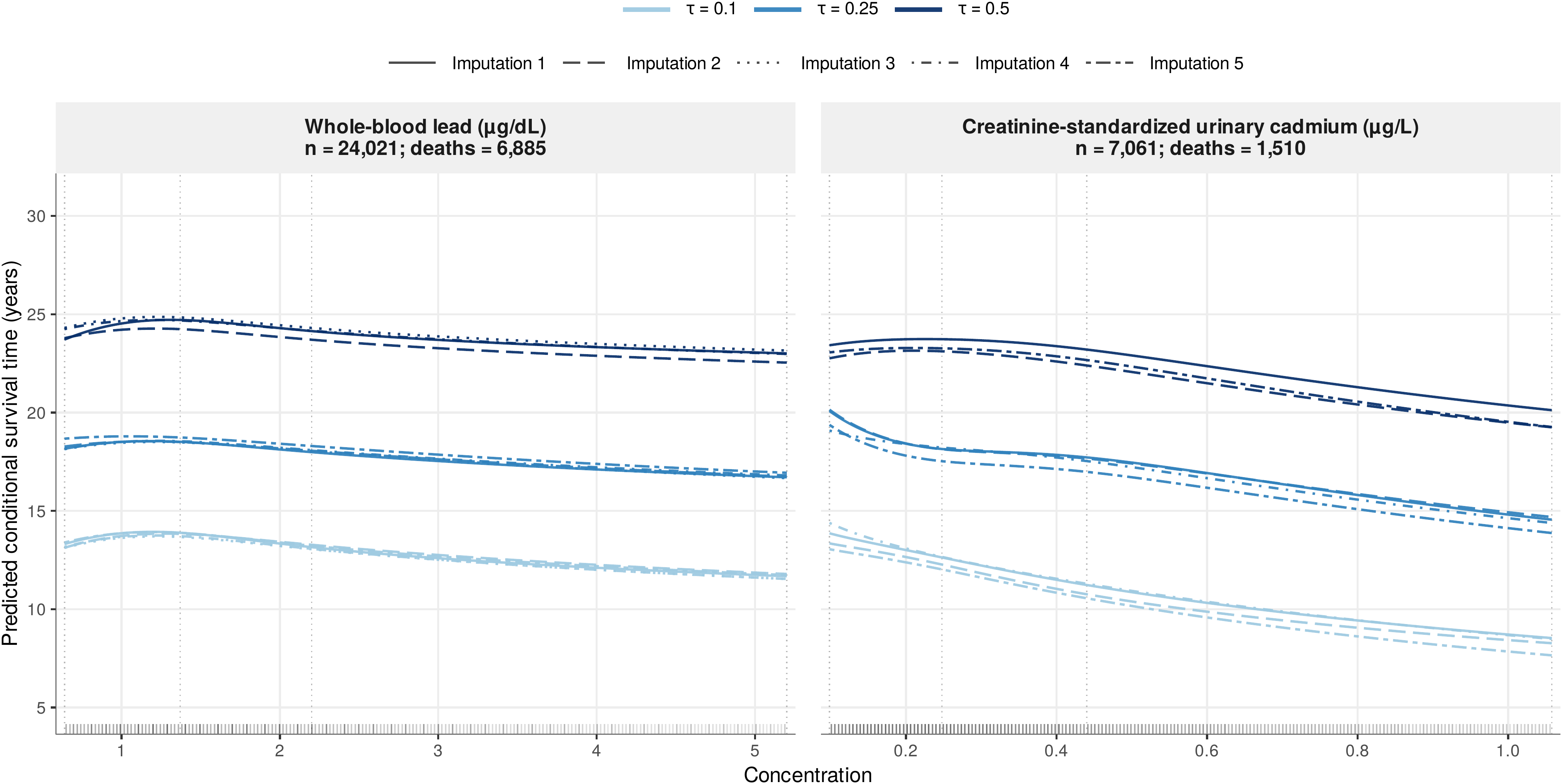
Blood lead and urinary cadmium concentrations and predicted conditional survival time. Predicted survival time at the 10th, 25th, and 50th conditional percentiles from censored quantile regression with a restricted cubic spline for log₂-transformed metal concentration (knots at the 5th, 35th, 65th, and 95th percentiles, dotted vertical lines), shown across the 5th–95th exposure percentiles. Blood lead: 24,021 participants and 6,885 deaths. Urinary cadmium: 7,061 participants and 1,510 deaths, restricted to eGFR ≥60 mL/min/1.73 m^2^. Other covariates were held at their median or mode. Colour indicates the survival-time percentile and line type the imputation; rug marks show the exposure distribution. Curves that were not estimable (cadmium, imputation 3; cadmium median, imputation 4; lead median, imputation 5) are not shown. eGFR=estimated glomerular filtration rate.

For whole-blood lead, predicted survival time was flat or rose slightly at the lowest concentrations and declined steadily above about 1.5 µg/dL, with the same pattern at τ = 0.25 and τ = 0.50 (figure 2).

### Sensitivity analyses

The cadmium and all-cause mortality association was robust to all four sensitivity analyses, whereas the lead and mortality association was robust to three but not to mutual adjustment for cadmium (appendix pp 19–23, 25). Portnoy estimates differed from the primary estimates by at most 0.13 years. Among participants aged 55 years or older, associations at τ = 0.10 were attenuated but persisted for lead (β −0.26 to −0.31 years; n = 14,735; 95% CIs excluded zero in four of five imputations) and cadmium (β −1.10 to −1.33 years; n = 3,982; all 95% CIs excluded zero). With mutual adjustment (n = 7,057; 1,510 deaths), lead was no longer negatively associated with survival time (β at τ = 0.10 0.59 to 0.69 years; all 95% CIs included zero), whereas cadmium remained so (β −1.78 to −2.03 years; all 95% CIs excluded zero). In the post-hoc analysis, replacing fish servings with dietary EPA plus DHA intake left the mercury and morality association unchanged (β 0.27 to 0.33 years at τ = 0.10).

### Nutrient effect modification

The only significant interaction was between blood mercury and serum α-carotene (p = 0.009), which did not remain significant after false discovery rate correction (q = 0.11; appendix pp 5, 12 –14).

### Replication in NHANES III

In NHANES III (1988–1994), a doubling of blood lead (n = 9,388; 6,146 deaths) was associated with 1.01–1.14 fewer years of survival time at τ = 0.10 and 0.81–1.26 fewer years across the three quantiles, and a doubling of urinary cadmium among participants with eGFR of 60 mL/min per 1.73 mZ or higher (n = 7,924; 4,823 deaths) with 1.38–1.42 fewer years at τ = 0.10 and 1.38–1.76 fewer years across the three quantiles. All estimates were statistically significant (95% CIs excluded zero, appendix pp 6, 15). Unlike in the primary cohort, the cadmium association in NHANES III was largest at the median (1.66–1.76 fewer years).

### Cancer and cardiovascular mortality

Of the 6,885 deaths in the blood lead cohort, 1,503 were from cancer and 2,233 from cardiovascular disease (appendix p 16). Cancer mortality was estimable only at τ = 0.10, and cardiovascular mortality at τ = 0.10 and, for lead and mercury, τ = 0.25. For cancer mortality, a doubling of urinary cadmium was associated with 2.02–2.34 fewer years at τ = 0.10 (391 cancer deaths; 95% CIs excluded zero in four of five imputations), compared with 1.71–1.92 fewer years for all-cause mortality. A doubling of blood lead was associated with 0.73–0.95 fewer years for cancer and 0.64–0.80 fewer years for cardiovascular mortality at τ = 0.10. Blood mercury estimates were positive for both cancer (0.49–0.58 years; 95% CIs included zero in four of five imputations) and cardiovascular mortality (0.50–0.55 years; four imputations; all 95% CIs excluded zero; appendix pp 17–18, 24).

## Discussion

In this prospective cohort of 24,338 US adults followed for up to 20 years, low-to-moderate urinary cadmium and whole-blood lead concentrations were associated with shorter survival time, with the largest absolute losses at the 10th percentile, where the earliest deaths occur. The cadmium association was robust to an alternative estimator, restriction to adults aged 55 years or older, and adjustment for lead, and was replicated in NHANES III; the lead association did not persist after adjustment for cadmium.

Previous NHANES studies of metals and mortality have mainly used Cox proportional harzards models.^6,7^ Other work has examined heterogeneous lead-associated survival in NHANES III using survival forests.^24^ Our study provided estimates in years and at lower survival distribution for earlier death. Compared with Cox models, our approach differs in three ways: CQR does not require proportional hazards; the estimand is expressed in years of survival time; and estimating effects at separate quantiles resolves associations in the lower tail of the survival distribution, where a given exposure effect translates into the largest absolute loss of life. A hazard ratio, on the other hand, compares instantaneous mortality rates among participants still alive, so changes in survivor composition can complicate their causal interpretation and cannot distinguish effect at lower vs. higher tail of the survival distribution.^11,12^

Our cadmium findings are consistent with previous NHANES and Strong Heart Study results. Comparing the 80th with the 20th percentile of urinary cadmium, Téllez-Plaza and colleagues reported hazard ratios of 1.52 (95% CI 1.00–2.29) for all-cause and 1.74 (1.07–2.83) for cardiovascular mortality in NHANES 1999–2004,^6^ and García-Esquinas and colleagues reported 1.30 (1.09–1.55) for cancer mortality in the Strong Heart Study.^25^ For the same contrast in blood lead, a recent Strong Heart Study analysis reported a hazard ratio of 1.15 (1.02–1.30) for cardiovascular mortality.^26^ In our cause-specific analyses, the cadmium association with cancer mortality was larger than with all-cause mortality, consistent with the classification of cadmium and cadmium compounds as a human carcinogen.^2^

An earlier NHANES study found no clear association between blood mercury and all-cause or cardiovascular mortality.^10^ Blood total mercury largely reflects organic mercury, predominantly methylmercury associated with fish and shellfish intake.^18^ Fish provides long-chain omega-3 fatty acids and selenium.^19^ Residual confounding by diet could contribute to the observed mercury association.

The null arsenic finding may reflect the within-person variability of spot-urine arsenic relative to long-term exposure, which could attenuate estimates towards the null.

Biological mechanisms support the cadmium and lead’s associations with mortality. Cadmium can promote oxidative stress and vascular injury.^27^ Lead promotes hypertension and vascular dysfunction and accumulates in bone, from which it is mobilised in later life.^28^ The American Heart Association claims lead and cadmium as cardiovascular risk factors.^5^

Our study has several strengths. CQR estimates associations on an interpretable time scale and resolves effects in the lower tail of the survival distribution. The CQR assumption that event and censoring times are independent conditional on covariates is plausible because censoring was administrative, determined by the examination date and the Dec 31, 2019. The nationally representative cohort with up to 20 years of follow-up provides greater statistical power and exposure range than previous single-cycle analyses. We used covariate-adjusted creatinine standardization to address urinary dilution.^16^ Multiple imputation and a design-stage eGFR restriction to limit residual confounding and reverse causation.

This study has several limitations. First, each metal was measured once, so exposure is potentially misclassified relative to long-term body burden. However, biomarkers were measured before follow-up, this error is unlikely to depend on survival time and would therefore bias associations towards the null. Second, residual confounding cannot be excluded. Third, the median is close to the identifiability boundary set by censoring, so some τ = 0.50 fits were not estimable. Fourth, the eGFR restriction and adjustment may also block part of an effect of cadmium mediated through kidney injury, biasing cadmium estimates towards the null, and do not address non-renal pathways, such as subclinical disease that raises both measured cadmium and mortality risk. Fifth, uncertainty intervals were not adjusted for the complex survey design. Finally, the cancer and cardiovascular mortality analyses assume that competing deaths are independent of cause-specific survival time given covariates.

In conclusion, low-to-moderate cadmium exposure was associated with shorter survival among US adults, with the largest absolute losses among those who die earliest. Continued action to reduce cadmium exposure could benefit the adults most vulnerable to premature death.

## Contributors

XF and JS conceived the study. XF did the analyses and wrote the first draft. JS supervised the study and critically revised the manuscript. XF and JS accessed and verified the underlying data. All authors had full access to all the data in the study, approved the final version, and had final responsibility for the decision to submit for publication.

## Data sharing

NHANES data and the public-use NDI-linked mortality files are publicly available from the National Center for Health Statistics. The analytic code will be available at https://github.com/fangxin0922/Heavy-metal-exposure-and-all-cause-mortality-in-U.S.-adults on publication.

## Data Availability

All data produced are available online at https://www.cdc.gov/nchs/nhanes/index.html

