## Supplementary appendix for "Environmental toxic metal exposure and conditional mortality survival among US adults: a prospective cohort study"

**Primary analysis**

The continuous National Health and Nutrition Examination Survey (NHANES) analyses included adults aged ≥40 years enrolled through 2014, with mortality follow-up to Dec 31, 2019 (table S1). Lead and mercury were measured in blood. Cadmium and arsenic were measured in urine and standardised using covariate-adjusted creatinine concentrations. Eligibility for the primary cadmium and arsenic analyses required observed baseline estimated glomerular filtration rate (eGFR) ≥60 mL/min per 1.73 $m^2$ before imputation.

Five imputed datasets were generated for each cohort using multiple imputation by chained equations (MICE), with 20 iterations. Exposures, mortality status, follow-up time, survey cycle, and survey design variables were not imputed. Table S2 reports observed missingness before imputation, and table S3 reports the primary results separately for each imputation.

For whole-blood lead and mercury, we used the Mobile Examination Center examination weight (the four-year weight for 1999–2002) and, for 2013–2014, the blood-metal subsample weight. For urinary cadmium and arsenic, we used the urinary subsample weight (the four-year subsample weight for 1999–2002). Weights were divided by the number of pooled two-year cycles (eight for lead and cadmium, and six for mercury and arsenic), with four-year weights first multiplied by two, and were then normalised to the cohort mean.

The 95% CIs were estimated from 400 delete-d jackknife resamples per imputation.

**Literature review for the nutrient analyses**

To select candidate effect modifiers, we did a focused narrative review of human studies linking metal biomarkers, nutrient status, and subsequent health (table S4). In NHANES III, higher blood lead and urinary cadmium were associated cross-sectionally with lower circulating vitamin C and carotenoids.[1] Plasma ascorbate was negatively associated with subsequent mortality in EPIC-Norfolk,[2] although vitamin C supplementation did not reduce major cardiovascular events in the Physicians' Health Study II.[3] Low serum α-carotene was associated with mortality in NHANES III.[4] The CARET trial, which tested β-carotene combined with retinol rather than α-carotene, found adverse outcomes among smokers and asbestos-exposed workers.[5] Blood lead and cadmium were negatively associated with red blood cell (RBC) folate in a cross-sectional NHANES analysis,[6] and folic acid supplementation lowered blood arsenic in folate-deficient, arsenic-exposed Bangladeshi adults.[7] We therefore examined serum vitamin C, serum α-carotene, and RBC folate. The nutrient biomarkers and metals were measured cross-sectionally.

**Nutrient effect modification**

Vitamin C analyses were restricted to 2003–2006. α-Carotene analyses covered 2001–2006 for lead and cadmium and 2003–2006 for mercury and arsenic. Assay conversion was applied to 2003–2004 values. RBC folate analyses covered 1999–2014 for lead and cadmium and 2003–2014 for mercury and arsenic, with conversions recommended by the Centers for Disease Control and Prevention across assay changes. Each metal–biomarker pair used a separate complete-case sample without imputation (table S5).

The continuous biomarker was scaled as (value − weighted P25)/(weighted P75 − weighted P25). Models included $\log_2$ metal concentration, the scaled biomarker, their product, and the covariates from the corresponding primary model. The interaction contrast was the metal association at biomarker P75 minus that at P25. The primary survival quantile was 0.10. Benjamini–Hochberg adjustment used the planned family of 12 primary interactions.

Inference used 400 survey replicates formed by resampling primary sampling units within strata, with positive affine rescaling ($\kappa$=2), fourfold variance correction, replicate-mean centring, and survey-based t degrees of freedom. All effect-modification results are presented in tables S6–S8 and figure S2.

**Replication in NHANES III**

We replicated our primary analyses in NHANES III (enrolment 1988–1994), with mortality follow-up through Dec 31, 2019. The blood lead cohort included 9388 participants and 6146 deaths, without an eGFR restriction or eGFR covariate. The urinary cadmium cohort included 7924 participants and 4823 deaths, with observed baseline eGFR ≥60 mL/min per 1.73 $m^2$ and continuous eGFR adjustment. Mercury and arsenic were not included due to data availability.

Five imputed datasets were generated for each metal by MICE with 50 iterations. Models used the same estimator, resampling, and covariates as the primary analysis, with normalised examination weights. NHANES III categories for fewer than 12 drinks could not be classified exactly as lifetime abstainer or former drinker, education had three school-year groups, physical activity was binary, and survey phase replaced survey cycle for covariates. Differences in period, exposure distribution, follow-up, questionnaire definitions, and urinary standardisation limit direct comparison of magnitudes across cohorts (table S9 and figure S3).

**Cancer and cardiovascular mortality**

We used category 002 for cancer mortality. We combined the heart disease (001) and cerebrovascular disease, including stroke (005) for cardiovascular mortality. For each analysis, we used the full metal cohort and censored deaths from other causes at the time of death (table S10). The analyses used the same imputed datasets, exposures, weights, covariates, and resampling as the primary analysis.

**Dose–response analysis**

The spline models used the primary blood lead cohort (24021 participants, 6885 all-cause deaths) and the primary urinary cadmium cohort (7061 participants, 1510 all-cause deaths), and were fitted in each of the five imputed datasets with the same covariates and weights as the primary models. For prediction, other covariates were held at their median or modal category. Approximate pointwise 95% intervals for predicted conditional survival quantiles were derived from the jackknife covariance matrix (333–396 finite draws of the 400 requested across the estimable cadmium quantiles). They are not simultaneous or pooled across imputations.

Eleven of 15 cadmium curves were estimable: imputation 3 yielded no curves, and imputation 4 yielded no median curve. Across the four available imputations, predicted conditional survival time at the 10th percentile decreased from 13.05–14.39 years at the 5th-percentile cadmium concentration (0.098 µg/L) to 7.65–8.53 years at the 95th percentile (1.058 µg/L). The within-imputation decreases were 5.08–5.90 years. At the 25th survival-time percentile, the corresponding prediction ranges were 19.08–20.14 and 13.87–14.69 years. Across three available median curves, they were 22.76–23.42 and 19.25–20.12 years.

**Sensitivity analyses**

The Portnoy and age ≥55 years analyses were run for all four metals. Analyses of the urinary metals were restricted to participants with observed baseline eGFR ≥60 mL/min per 1.73 $m^2$, as in the primary analysis (table S13). Portnoy models used the same five imputed datasets, covariates, and weights as the primary analysis, so only the estimator differed. The cadmium and arsenic age ≥55 years cohorts and the mutual-adjustment cohort were imputed separately after the eligibility restrictions (five datasets, with 50 MICE iterations). The mutual-adjustment models included $\log_2$ lead and $\log_2$ cadmium as additive terms, with urinary cadmium sampling weights. The lead estimate comes from a model with the primary lead adjustment set, and the cadmium estimate from a model that also adjusted for eGFR. In a post-hoc analysis, fish intake in the mercury model was replaced with dietary EPA plus DHA intake (table S19). Imputation-specific results are given in tables S14–S16 and S19 and summarised in table S17.

**Figure S1. DAG**

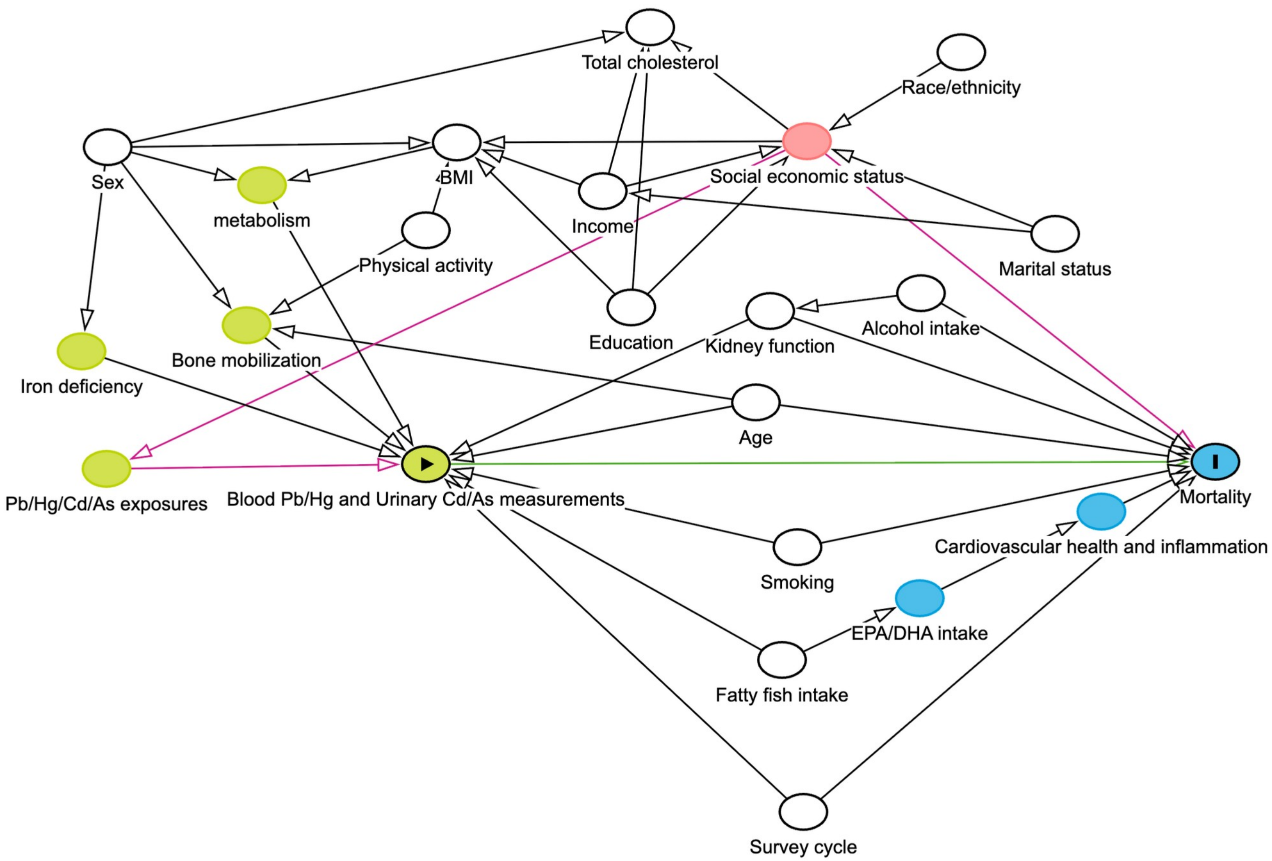

As=arsenic; BMI=body-mass index; Cd=cadmium; DHA=docosahexaenoic acid; EPA=eicosapentaenoic acid; Hg=mercury; Pb=lead.

**Figure S2. Primary nutrient effect-modification contrasts**

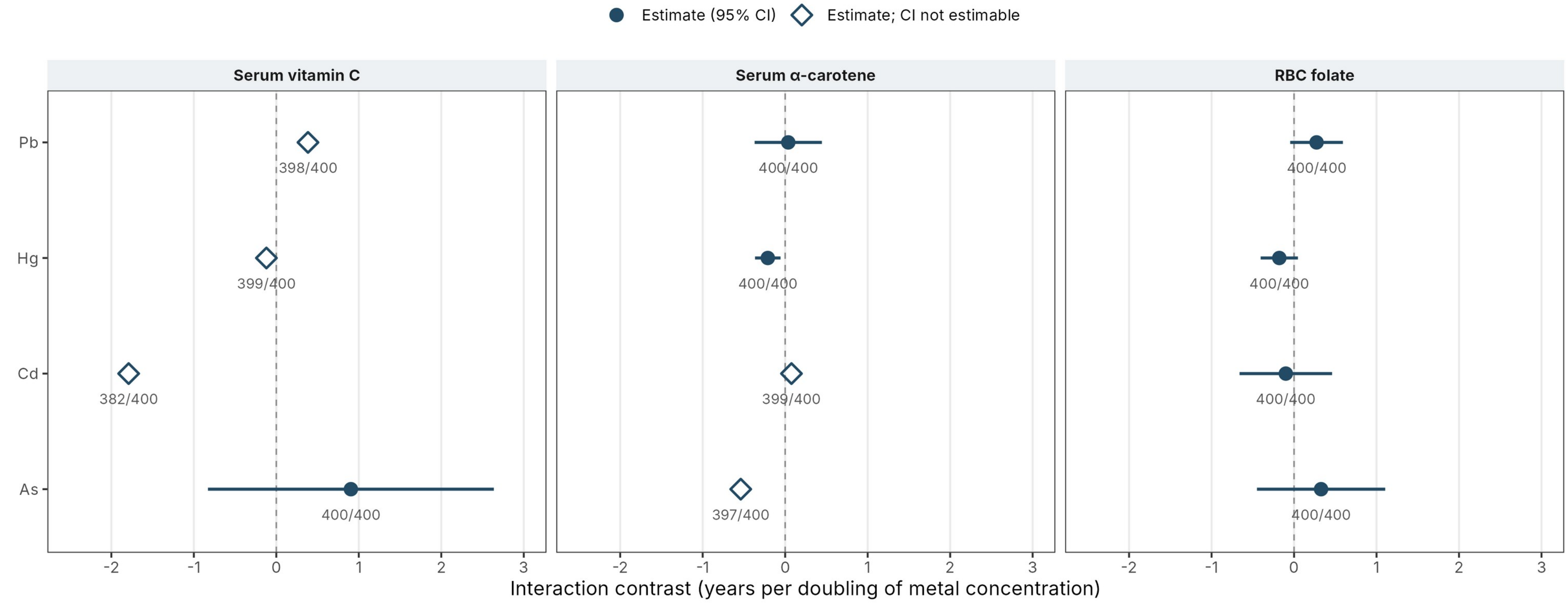

Interaction contrasts (difference in the per-doubling metal association at the 75th versus 25th biomarker percentile) at the 10th conditional percentile of survival time. Filled circles show estimates with 95% CIs. Open diamonds show estimates for which a CI could not be computed. No interaction met the Benjamini–Hochberg threshold (q<0.05).

As=arsenic; BH=Benjamini–Hochberg; Cd=cadmium; CI=confidence interval; Hg=mercury; Pb=lead; RBC=red blood cell.

**Figure S3. NHANES III results by imputation**

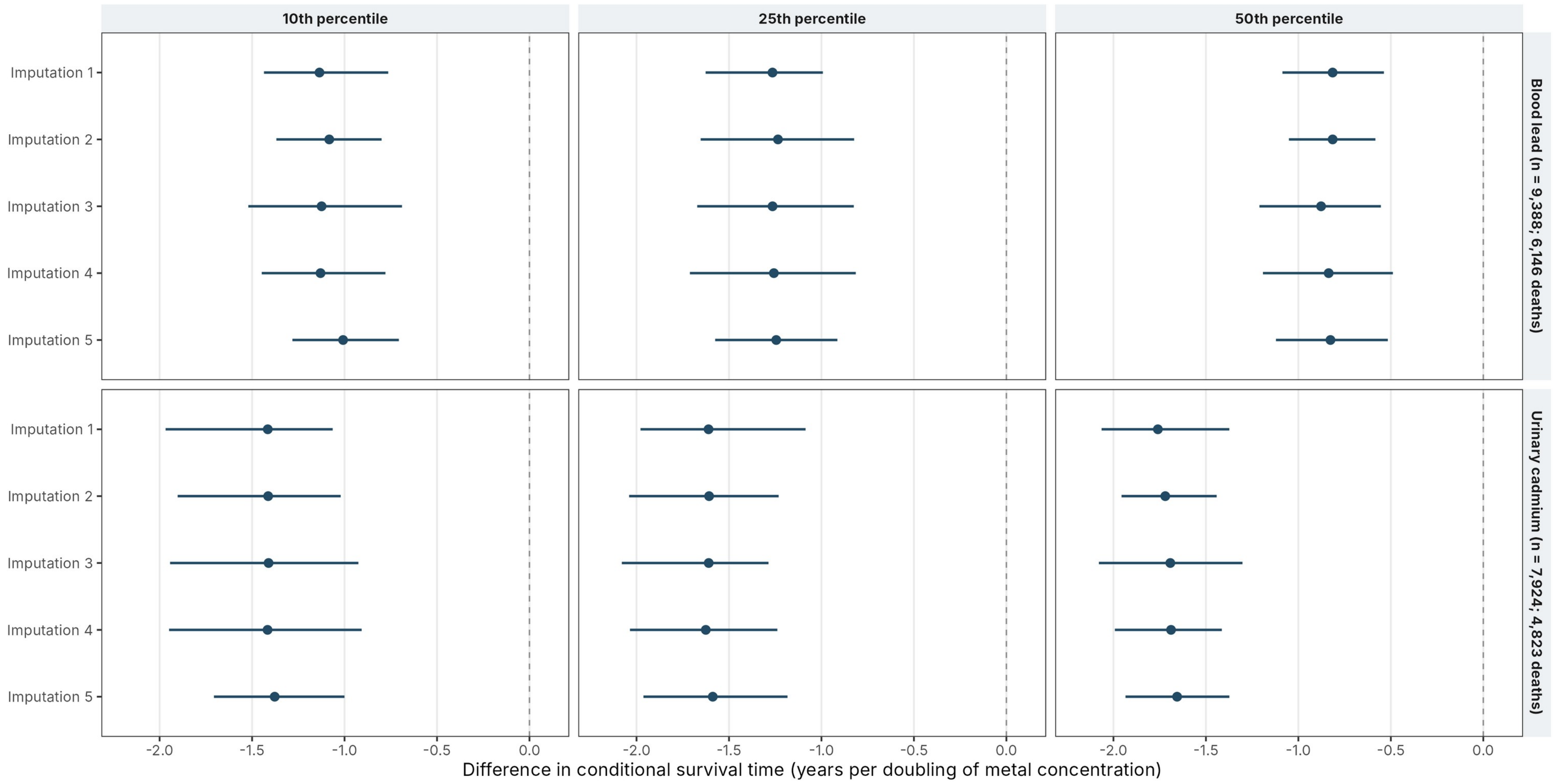

Estimates (95% CI) for blood lead and urinary cadmium at the 10th, 25th, and 50th conditional survival-time percentiles, shown separately for each imputation. The cadmium cohort was restricted to eGFR ≥60 mL/min per 1.73 m².

CI=confidence interval; eGFR=estimated glomerular filtration rate; NHANES=National Health and Nutrition Examination Survey.

**Table S1. Primary and replication cohorts and imputation specifications**

| Cohort | Metal | Enrolment | N / deaths | M / iterations | Available cells |
|---|---|---|---|---|---|
| **Continuous NHANES** | Pb | 1999–2014 | 24021 / 6885 | 5 / 20 | 13/15 |
| **Continuous NHANES** | Hg | 2003–2014 | 18258 / 4358 | 5 / 20 | 12/15 |
| **Continuous NHANES** | Cd | 1999–2014 | 7061 / 1510 | 5 / 20 | 15/15 |
| **Continuous NHANES** | As | 2003–2014 | 5552 / 960 | 5 / 20 | 12/15 |
| **NHANES III** | Pb | 1988–1994 | 9388 / 6146 | 5 / 50 | 15/15 |
| **NHANES III** | Cd | 1988–1994 | 7924 / 4823 | 5 / 50 | 15/15 |

Counts are unweighted. M=number of imputations. Available cells=number of estimable results across five imputations and three quantiles. All cohorts comprise adults aged ≥40 years with mortality follow-up through Dec 31, 2019. eGFR is in mL/min per 1.73 m².

As=arsenic; Cd=cadmium; eGFR=estimated glomerular filtration rate; Hg=mercury; NHANES=National Health and Nutrition Examination Survey; Pb=lead.

**Table S2. Observed covariate missingness in the primary cohorts**

| Covariate | Pb (N=24021) | Hg (N=18258) | Cd (N=7061) | As (N=5552) |
|---|---|---|---|---|
| **Age** | 0 (0.0) | 0 (0.0) | 0 (0.0) | 0 (0.0) |
| **Sex** | 0 (0.0) | 0 (0.0) | 0 (0.0) | 0 (0.0) |
| **Race and ethnicity** | 0 (0.0) | 0 (0.0) | 0 (0.0) | 0 (0.0) |
| **Education** | 45 (0.1) | 29 (0.1) | 6 (0.0) | 3 (0.0) |
| **Marital status** | 297 (1.3) | 12 (0.1) | 84 (1.2) | 2 (0.1) |
| **Income-to-poverty ratio** | 2084 (7.0) | 1455 (6.4) | 586 (6.7) | 422 (6.1) |
| **BMI** | 566 (1.8) | 321 (1.3) | 0 (0.0) | 0 (0.0) |
| **Smoking pack-years** | 1395 (5.6) | 917 (5.1) | 390 (5.6) | 258 (5.0) |
| **Alcohol** | 1772 (6.6) | 1475 (7.2) | 439 (5.5) | 395 (6.4) |
| **Physical activity** | 490 (1.6) | 261 (1.2) | 127 (1.5) | 66 (1.0) |
| **Survey cycle** | 0 (0.0) | 0 (0.0) | 0 (0.0) | 0 (0.0) |
| **Total cholesterol** | 406 (1.4) | 265 (1.2) | 3 (0.1) | 0 (0.0) |
| **eGFR** | 472 (1.6) | 339 (1.5) | 0 (0.0) | 0 (0.0) |
| **Fish intake** | 1244 (4.7) | 996 (4.9) | 332 (3.9) | 293 (4.5) |

Data are n missing (survey-weighted %) before imputation. Exposures, mortality, and follow-up time were complete by design. eGFR was an adjustment variable only in the cadmium and arsenic models, and fish intake only in the mercury model.

As=arsenic; BMI=body-mass index; Cd=cadmium; eGFR=estimated glomerular filtration rate; Hg=mercury; Pb=lead.

**Table S3. Primary CQR results by imputation**

| Metal | 10th percentile | 25th percentile | 50th percentile |
|---|---|---|---|
| **Blood lead (Pb)**<br>**N=24021; deaths=6885** | I1: −0.671 (−0.906, −0.411); p<0.0001<br>I2: −0.685 (−0.923, −0.437); p<0.0001<br>I3: −0.679 (−0.995, −0.354); p<0.0001<br>I4: −0.706 (−0.974, −0.461); p<0.0001<br>I5: −0.625 (−0.890, −0.310); p<0.0001 | I1: −0.555 (−0.916, −0.140); p=0.0051<br>I2: −0.607 (−0.813, −0.380); p<0.0001<br>I3: −0.587 (−0.817, −0.365); p<0.0001<br>I4: −0.650 (−0.895, −0.388); p<0.0001<br>I5: −0.612 (−0.887, −0.268); p=0.0001 | I1: NE<br>I2: −0.613 (−0.815, −0.383); p<0.0001<br>I3: −0.626 (−0.879, −0.281); p<0.0001<br>I4: NE<br>I5: −0.620 (−0.913, −0.319); p<0.0001 |
| **Blood mercury (Hg)**<br>**N=18258; deaths=4358** | I1: 0.328 (0.168, 0.487); p<0.0001<br>I2: 0.286 (0.172, 0.420); p<0.0001<br>I3: 0.327 (0.160, 0.483); p<0.0001<br>I4: 0.287 (0.098, 0.456); p=0.0016<br>I5: 0.327 (0.165, 0.524); p=0.0003 | I1: 0.477 (0.320, 0.600); p<0.0001<br>I2: NE<br>I3: 0.511 (0.312, 0.704); p<0.0001<br>I4: 0.502 (0.307, 0.689); p<0.0001<br>I5: 0.529 (0.315, 0.813); p<0.0001 | I1: 0.572 (0.336, 0.797); p<0.0001<br>I2: NE<br>I3: NE<br>I4: 0.579 (0.378, 0.776); p<0.0001<br>I5: 0.538 (0.275, 0.714); p<0.0001 |
| **Urinary cadmium (Cd)**<br>**N=7061; deaths=1510** | I1: −1.763 (−2.514, −1.008); p<0.0001<br>I2: −1.708 (−2.535, −0.962); p<0.0001<br>I3: −1.723 (−2.480, −1.032); p<0.0001<br>I4: −1.741 (−2.365, −1.131); p<0.0001<br>I5: −1.919 (−2.534, −1.140); p<0.0001 | I1: −1.638 (−2.117, −1.117); p<0.0001<br>I2: −1.571 (−2.134, −0.908); p<0.0001<br>I3: −1.501 (−1.963, −0.865); p<0.0001<br>I4: −1.632 (−2.209, −1.070); p<0.0001<br>I5: −1.665 (−2.090, −1.025); p<0.0001 | I1: −1.347 (−1.938, −0.755); p<0.0001<br>I2: −1.521 (−2.135, −0.890); p<0.0001<br>I3: −1.508 (−2.103, −0.894); p<0.0001<br>I4: −1.535 (−2.234, −0.945); p<0.0001<br>I5: −1.395 (−2.375, −0.628); p=0.0017 |
| **Urinary arsenic (As)**<br>**N=5552; deaths=960** | I1: 0.177 (−0.379, 0.772); p=0.5468<br>I2: 0.321 (−0.430, 1.089); p=0.4070<br>I3: 0.058 (−0.655, 1.049); p=0.8941<br>I4: 0.075 (−0.624, 0.792); p=0.8364<br>I5: 0.035 (−0.540, 0.603); p=0.9035 | I1: 0.327 (−0.775, 1.151); p=0.5058<br>I2: 0.416 (−0.293, 1.220); p=0.2818<br>I3: 0.541 (−0.183, 1.264); p=0.1426<br>I4: 0.536 (−0.205, 1.349); p=0.1763<br>I5: 0.466 (−0.644, 1.369); p=0.3642 | I1: NE<br>I2: 0.289 (−0.780, 1.368); p=0.5979<br>I3: 0.300 (−0.341, 1.056); p=0.3994<br>I4: NE<br>I5: NE |

Values are the difference in the conditional quantile of survival time (years) per doubling of metal concentration (95% CI) and p value for each imputation (I1–I5). NE=not estimable. 95% CIs were estimated from 400 delete-d jackknife resamples.

Models were adjusted for age, age², sex, race and ethnicity, education, marital status, income-to-poverty ratio, BMI, pack-years, alcohol use, physical activity, survey cycle, and total cholesterol. Cadmium and arsenic models were additionally adjusted for eGFR and restricted to eGFR ≥60 mL/min per 1.73 m². The mercury model was additionally adjusted for fish intake.

As=arsenic; BMI=body-mass index; Cd=cadmium; CQR=censored quantile regression; eGFR=estimated glomerular filtration rate; Hg=mercury; Pb=lead.

**Table S4. Human evidence informing the selected nutrient analyses**

| Biomarker / context | Study and design | Evidence |
|---|---|---|
| **Vitamin C / carotenoids** | Lee et al, 2006;[1] NHANES III cross-sectional | Higher blood Pb and urinary Cd were associated with lower antioxidant biomarker concentrations. |
| **Vitamin C** | Khaw et al, 2001;[2] prospective EPIC-Norfolk cohort | Higher plasma ascorbate was associated with lower subsequent mortality. |
| **Vitamin C** | Sesso et al, 2008;[3] randomised Physicians' Health Study II | Vitamin C supplementation did not reduce major cardiovascular events. |
| **α-Carotene** | Shardell et al, 2011;[4] NHANES III mortality follow-up | Low serum α-carotene and carotenoid combinations were associated with mortality. |
| **Carotenoid intervention context** | Omenn et al, 1996;[5] randomised CARET trial | β-Carotene plus retinol produced adverse lung-cancer and mortality outcomes in high-risk participants. |
| **RBC folate** | Wang and Chen, 2022;[6] NHANES cross-sectional | Blood Pb and Cd were negatively associated with RBC folate. |
| **Folate / arsenic metabolism** | Gamble et al, 2007;[7] randomised trial | Folic acid lowered blood arsenic in folate-deficient, exposed Bangladeshi adults. |

Cd=cadmium; NHANES=National Health and Nutrition Examination Survey; Pb=lead; RBC=red blood cell.

**Table S5. Effect-modification cohorts and biomarker reference levels**

| Metal | Biomarker | Enrolment | N | Deaths | Units | P25 | P75 |
|---|---|---|---|---|---|---|---|
| **Blood lead (Pb)** | Serum vitamin C | 2003–2006 | 4465 | 1499 | mg/dL | 0.600 | 1.290 |
| **Blood mercury (Hg)** | Serum vitamin C | 2003–2006 | 4400 | 1477 | mg/dL | 0.600 | 1.290 |
| **Urinary cadmium (Cd)** | Serum vitamin C | 2003–2006 | 1312 | 354 | mg/dL | 0.560 | 1.320 |
| **Urinary arsenic (As)** | Serum vitamin C | 2003–2006 | 1315 | 354 | mg/dL | 0.560 | 1.320 |
| **Blood lead (Pb)** | Serum α-carotene | 2001–2006 | 6666 | 2294 | μg/dL | 1.660 | 5.663 |
| **Blood mercury (Hg)** | Serum α-carotene | 2003–2006 | 4404 | 1479 | μg/dL | 1.611 | 5.644 |
| **Urinary cadmium (Cd)** | Serum α-carotene | 2001–2006 | 1942 | 560 | μg/dL | 1.600 | 5.831 |
| **Urinary arsenic (As)** | Serum α-carotene | 2003–2006 | 1315 | 355 | μg/dL | 1.591 | 5.693 |
| **Blood lead (Pb)** | RBC folate | 1999–2014 | 17936 | 4776 | nmol/L | 928.000 | 1592.653 |
| **Blood mercury (Hg)** | RBC folate | 2003–2014 | 13709 | 3134 | nmol/L | 914.954 | 1580.000 |
| **Urinary cadmium (Cd)** | RBC folate | 1999–2014 | 5570 | 1147 | nmol/L | 933.000 | 1560.000 |
| **Urinary arsenic (As)** | RBC folate | 2003–2014 | 4485 | 770 | nmol/L | 920.611 | 1550.517 |

Counts are unweighted complete cases. P25 and P75 are survey-weighted biomarker percentiles used as evaluation levels. Urinary cadmium and arsenic cohorts were restricted to eGFR ≥60 mL/min per 1.73 $m^2$.

As=arsenic; Cd=cadmium; eGFR=estimated glomerular filtration rate; Hg=mercury; Pb=lead; RBC=red blood cell.

**Table S6. Effect modification by serum vitamin C**

| Metal | 10th percentile | 25th percentile | 50th percentile |
|---|---|---|---|
| **Blood lead (Pb)<br>N=4465; deaths=1499** | P25: −0.531 (CI unavailable)<br>P75: −0.147 (CI unavailable)<br>IC: 0.384 (CI unavailable)<br>p=NE; BH q=NE<br>Finite IC replicates: 398/400 | P25: −0.324 (CI unavailable)<br>P75: 0.003 (CI unavailable)<br>IC: 0.327 (CI unavailable)<br>p=NE<br>Finite IC replicates: 396/400 | P25: −0.031 (CI unavailable)<br>P75: 0.081 (CI unavailable)<br>IC: 0.112 (CI unavailable)<br>p=NE<br>Finite IC replicates: 381/400 |
| **Blood mercury (Hg)<br>N=4400; deaths=1477** | P25: 0.315 (−0.108, 0.739)<br>P75: 0.196 (CI unavailable)<br>IC: −0.120 (CI unavailable)<br>p=NE; BH q=NE<br>Finite IC replicates: 399/400 | P25: 0.504 (CI unavailable)<br>P75: 0.216 (CI unavailable)<br>IC: −0.289 (−0.729, 0.152)<br>p=0.1907<br>Finite IC replicates: 400/400 | P25: 0.859 (CI unavailable)<br>P75: 0.437 (CI unavailable)<br>IC: −0.422 (CI unavailable)<br>p=NE<br>Finite IC replicates: 388/400 |
| **Urinary cadmium (Cd)<br>N=1312; deaths=354** | P25: −0.795 (CI unavailable)<br>P75: −2.585 (CI unavailable)<br>IC: −1.790 (CI unavailable)<br>p=NE; BH q=NE<br>Finite IC replicates: 382/400 | P25: NE<br>P75: NE<br>IC: NE<br>p=NE<br>Finite IC replicates: 167/400 | P25: NE<br>P75: NE<br>IC: NE<br>p=NE<br>Finite IC replicates: 2/400 |
| **Urinary arsenic (As)<br>N=1315; deaths=354** | P25: 0.282 (−1.069, 1.634)<br>P75: 1.187 (−0.604, 2.977)<br>IC: 0.904 (−0.829, 2.637)<br>p=0.2951; BH q=0.8853<br>Finite IC replicates: 400/400 | P25: −0.591 (CI unavailable)<br>P75: 0.151 (CI unavailable)<br>IC: 0.742 (CI unavailable)<br>p=NE<br>Finite IC replicates: 300/400 | P25: NE<br>P75: NE<br>IC: NE<br>p=NE<br>Finite IC replicates: 63/400 |

P25 and P75 are the metal associations (years per doubling, with 95% CIs) at the 25th and 75th biomarker percentiles. IC is the interaction contrast (P75 minus P25). p values test IC=0. Benjamini–Hochberg (BH) q values are given for the 10th percentile across the 12 primary tests. A CI is reported only when all 400 survey replicates gave a finite estimate. “Finite IC replicates” gives the number of finite interaction contrasts. Models used complete cases (table S5) and the primary adjustment set plus the biomarker and its product with the metal. NE=not estimable.

As=arsenic; BH=Benjamini–Hochberg; Cd=cadmium; Hg=mercury; Pb=lead.

**Table S7. Effect modification by serum α-carotene**

| Metal | 10th percentile | 25th percentile | 50th percentile |
|---|---|---|---|
| **Blood lead (Pb)**<br>**N=6666; deaths=2294** | P25: −0.604 (−1.344, 0.137)<br>P75: −0.566 (−1.307, 0.175)<br>IC: 0.037 (−0.370, 0.445)<br>p=0.8543; BH q=1.0000<br>Finite IC replicates: 400/400 | P25: −0.474 (CI unavailable)<br>P75: −0.482 (CI unavailable)<br>IC: −0.008 (CI unavailable)<br>p=NE<br>Finite IC replicates: 395/400 | P25: −0.696 (CI unavailable)<br>P75: −0.339 (CI unavailable)<br>IC: 0.357 (CI unavailable)<br>p=NE<br>Finite IC replicates: 387/400 |
| **Blood mercury (Hg)**<br>**N=4404; deaths=1479** | P25: 0.453 (0.007, 0.899)<br>P75: 0.242 (−0.167, 0.651)<br>IC: −0.211 (−0.365, −0.056)<br>p=0.0092; BH q=0.1108<br>Finite IC replicates: 400/400 | P25: 0.520 (CI unavailable)<br>P75: 0.281 (CI unavailable)<br>IC: −0.239 (CI unavailable)<br>p=NE<br>Finite IC replicates: 398/400 | P25: 0.573 (CI unavailable)<br>P75: 0.530 (CI unavailable)<br>IC: −0.043 (CI unavailable)<br>p=NE<br>Finite IC replicates: 386/400 |
| **Urinary cadmium (Cd)**<br>**N=1942; deaths=560** | P25: −2.343 (CI unavailable)<br>P75: −2.267 (CI unavailable)<br>IC: 0.076 (CI unavailable)<br>p=NE; BH q=NE<br>Finite IC replicates: 399/400 | P25: −1.715 (−3.169, −0.260)<br>P75: −1.344 (−2.326, −0.363)<br>IC: 0.371 (−0.472, 1.213)<br>p=0.3806<br>Finite IC replicates: 400/400 | P25: −1.714 (CI unavailable)<br>P75: −1.731 (CI unavailable)<br>IC: −0.017 (CI unavailable)<br>p=NE<br>Finite IC replicates: 389/400 |
| **Urinary arsenic (As)**<br>**N=1315; deaths=355** | P25: 0.956 (CI unavailable)<br>P75: 0.417 (CI unavailable)<br>IC: −0.539 (CI unavailable)<br>p=NE; BH q=NE<br>Finite IC replicates: 397/400 | P25: 0.242 (CI unavailable)<br>P75: −0.285 (CI unavailable)<br>IC: −0.528 (CI unavailable)<br>p=NE<br>Finite IC replicates: 259/400 | P25: NE<br>P75: NE<br>IC: NE<br>p=NE<br>Finite IC replicates: 81/400 |

P25 and P75 are the metal associations (years per doubling, with 95% CIs) at the 25th and 75th biomarker percentiles. IC is the interaction contrast (P75 minus P25). p values test IC=0. Benjamini–Hochberg (BH) q values are given for the 10th percentile across the 12 primary tests. A CI is reported only when all 400 survey replicates gave a finite estimate. “Finite IC replicates” gives the number of finite interaction contrasts. Models used complete cases (table S5) and the primary adjustment set plus the biomarker and its product term with the metal. NE=not estimable.

As=arsenic; BH=Benjamini–Hochberg; Cd=cadmium; Hg=mercury; Pb=lead.

### Table S8. Effect modification by RBC folate

| Metal | 10th percentile | 25th percentile | 50th percentile |
|---|---|---|---|
| **Blood lead (Pb)**<br>**N=17936; deaths=4776** | P25: −0.960 (−1.386, −0.533)<br>P75: −0.687 (−1.067, −0.307)<br>IC: 0.273 (−0.046, 0.592)<br>p=0.0933; BH q=0.4761<br>Finite IC replicates: 400/400 | P25: −0.864 (CI unavailable)<br>P75: −0.761 (CI unavailable)<br>IC: 0.103 (CI unavailable)<br>p=NE<br>Finite IC replicates: 397/400 | P25: −0.740 (CI unavailable)<br>P75: −0.572 (CI unavailable)<br>IC: 0.167 (CI unavailable)<br>p=NE<br>Finite IC replicates: 388/400 |
| **Blood mercury (Hg)**<br>**N=13709; deaths=3134** | P25: 0.404 (0.097, 0.710)<br>P75: 0.225 (0.010, 0.439)<br>IC: −0.179 (−0.405, 0.047)<br>p=0.1190; BH q=0.4761<br>Finite IC replicates: 400/400 | P25: 0.687 (0.379, 0.994)<br>P75: 0.608 (0.376, 0.840)<br>IC: −0.079 (−0.312, 0.154)<br>p=0.5043<br>Finite IC replicates: 400/400 | P25: 0.567 (CI unavailable)<br>P75: 0.576 (CI unavailable)<br>IC: 0.009 (CI unavailable)<br>p=NE<br>Finite IC replicates: 394/400 |
| **Urinary cadmium (Cd)**<br>**N=5570; deaths=1147** | P25: −1.855 (−2.765, −0.945)<br>P75: −1.955 (−2.948, −0.961)<br>IC: −0.100 (−0.661, 0.461)<br>p=0.7250; BH q=1.0000<br>Finite IC replicates: 400/400 | P25: −1.667 (CI unavailable)<br>P75: −1.559 (CI unavailable)<br>IC: 0.108 (CI unavailable)<br>p=NE<br>Finite IC replicates: 394/400 | P25: −1.379 (CI unavailable)<br>P75: −1.521 (CI unavailable)<br>IC: −0.142 (CI unavailable)<br>p=NE<br>Finite IC replicates: 376/400 |
| **Urinary arsenic (As)**<br>**N=4485; deaths=770** | P25: 0.087 (−1.198, 1.372)<br>P75: 0.415 (−0.588, 1.417)<br>IC: 0.328 (−0.450, 1.105)<br>p=0.4049; BH q=0.9717<br>Finite IC replicates: 400/400 | P25: 0.354 (CI unavailable)<br>P75: 0.551 (CI unavailable)<br>IC: 0.197 (CI unavailable)<br>p=NE<br>Finite IC replicates: 396/400 | P25: 0.140 (CI unavailable)<br>P75: 0.236 (CI unavailable)<br>IC: 0.096 (CI unavailable)<br>p=NE<br>Finite IC replicates: 316/400 |

P25 and P75 are the metal associations (years per doubling, with 95% CIs) at the 25th and 75th biomarker percentiles. IC is the interaction contrast (P75 minus P25). p values test IC=0. Benjamini–Hochberg (BH) q values are given for the 10th percentile across the 12 primary tests. A CI is reported only when all 400 survey replicates gave a finite estimate. “Finite IC replicates” gives the number of finite interaction contrasts. Models used complete cases (table S5) and the primary adjustment set plus the biomarker and its product with the metal. NE=not estimable.

As=arsenic; BH=Benjamini–Hochberg; Cd=cadmium; Hg=mercury; Pb=lead; RBC=red blood cell.

**Table S9. NHANES III CQR results by imputation**

| Metal | 10th percentile | 25th percentile | 50th percentile |
|---|---|---|---|
| **Blood lead**<br>**N=9388; deaths=6146** | I1: −1.136 (−1.436, −0.764); p<0.0001<br>I2: −1.083 (−1.369, −0.800); p<0.0001<br>I3: −1.124 (−1.520, −0.689); p<0.0001<br>I4: −1.130 (−1.448, −0.779); p<0.0001<br>I5: −1.008 (−1.282, −0.706); p<0.0001 | I1: −1.265 (−1.626, −0.992); p<0.0001<br>I2: −1.235 (−1.653, −0.823); p<0.0001<br>I3: −1.264 (−1.672, −0.825); p<0.0001<br>I4: −1.257 (−1.711, −0.814); p<0.0001<br>I5: −1.245 (−1.575, −0.914); p<0.0001 | I1: −0.815 (−1.086, −0.538); p<0.0001<br>I2: −0.815 (−1.051, −0.583); p<0.0001<br>I3: −0.877 (−1.210, −0.554); p<0.0001<br>I4: −0.836 (−1.191, −0.489); p<0.0001<br>I5: −0.827 (−1.121, −0.516); p<0.0001 |
| **Urinary cadmium**<br>**N=7924; deaths=4823** | I1: −1.415 (−1.968, −1.064); p<0.0001<br>I2: −1.413 (−1.902, −1.021); p<0.0001<br>I3: −1.411 (−1.943, −0.925); p<0.0001<br>I4: −1.416 (−1.949, −0.907); p<0.0001<br>I5: −1.378 (−1.706, −1.001); p<0.0001 | I1: −1.611 (−1.979, −1.086); p<0.0001<br>I2: −1.608 (−2.040, −1.232); p<0.0001<br>I3: −1.610 (−2.079, −1.286); p<0.0001<br>I4: −1.625 (−2.035, −1.239); p<0.0001<br>I5: −1.588 (−1.963, −1.184); p<0.0001 | I1: −1.760 (−2.064, −1.373); p<0.0001<br>I2: −1.720 (−1.956, −1.442); p<0.0001<br>I3: −1.692 (−2.079, −1.302); p<0.0001<br>I4: −1.689 (−1.992, −1.414); p<0.0001<br>I5: −1.656 (−1.935, −1.373); p<0.0001 |

Values are the difference in the conditional quantile of survival time (years) per doubling of metal concentration (95% CI) and p value for each imputation (I1–I5). 95% CIs were estimated from 400 delete-d jackknife resamples.

The cadmium model was restricted to eGFR ≥60 mL/min per 1.73 m² and adjusted for eGFR. Models were adjusted for age, age², sex, race and ethnicity, education, marital status, poverty-income ratio, BMI, pack-years, alcohol use, physical activity, survey phase, and total cholesterol.

BMI=body-mass index; CQR=censored quantile regression; eGFR=estimated glomerular filtration rate; NHANES=National Health and Nutrition Examination Survey.

### Table S10. Cohorts and underlying causes of death for the cancer and cardiovascular mortality analyses

| Metal / enrolment | N | All deaths | Cancer | Heart disease | Stroke | Cardiovascular | Other causes | Alive |
|---|---|---|---|---|---|---|---|---|
| **Blood lead (Pb) 1999–2014** | 24021 | 6885 | 1503 | 1812 | 421 | 2233 | 3149 | 17136 |
| **Blood mercury (Hg) 2003–2014** | 18258 | 4358 | 987 | 1146 | 264 | 1410 | 1961 | 13900 |
| **Urinary cadmium (Cd) 1999–2014** | 7061 | 1510 | 391 | 370 | 78 | 448 | 671 | 5551 |
| **Urinary arsenic (As) 2003–2014** | 5552 | 960 | 264 | 232 | 51 | 283 | 413 | 4592 |

Counts are unweighted. Cohorts overlap. Cancer: underlying-cause category 002. Cardiovascular: heart disease (001) and cerebrovascular disease (005). Other causes exclude cancer and cardiovascular deaths. Urinary cadmium and arsenic cohorts were restricted to eGFR ≥60 mL/min per 1.73 $m^2$.

As=arsenic; Cd=cadmium; eGFR=estimated glomerular filtration rate; Hg=mercury; Pb=lead.

### Table S11. Cancer mortality: cause-censored CQR results by imputation

| Metal | 10th percentile | 25th percentile | 50th percentile |
|---|---|---|---|
| **Blood lead (Pb)**<br>**N=24021**<br>**Cancer deaths=1503** | I1: −0.786 (−1.626, −0.050); p=0.0508<br>I2: −0.947 (−1.707, −0.326); p=0.0072<br>I3: −0.799 (−1.533, 0.024); p=0.0443<br>I4: −0.726 (−1.484, −0.020); p=0.0518<br>I5: −0.800 (−1.390, −0.234); p=0.0067 | I1: NE<br>I2: NE<br>I3: NE<br>I4: NE<br>I5: NE | I1: NE<br>I2: NE<br>I3: NE<br>I4: NE<br>I5: NE |
| **Blood mercury (Hg)**<br>**N=18258**<br>**Cancer deaths=987** | I1: 0.544 (−0.051, 1.354); p=0.1293<br>I2: 0.517 (−0.315, 1.327); p=0.2175<br>I3: 0.541 (−0.338, 1.491); p=0.2464<br>I4: 0.493 (−0.035, 0.935); p=0.0461<br>I5: 0.580 (0.209, 0.939); p=0.0018 | I1: NE<br>I2: NE<br>I3: NE<br>I4: NE<br>I5: NE | I1: NE<br>I2: NE<br>I3: NE<br>I4: NE<br>I5: NE |
| **Urinary cadmium (Cd)**<br>**N=7061**<br>**Cancer deaths=391** | I1: −2.250 (−4.499, −0.466); p=0.0287<br>I2: −2.174 (−4.139, −0.157); p=0.0324<br>I3: −2.308 (−4.424, 0.041); p=0.0427<br>I4: −2.017 (−3.818, −0.354); p=0.0225<br>I5: −2.339 (−3.957, −0.899); p=0.0027 | I1: NE<br>I2: NE<br>I3: NE<br>I4: NE<br>I5: NE | I1: NE<br>I2: NE<br>I3: NE<br>I4: NE<br>I5: NE |
| **Urinary arsenic (As)**<br>**N=5552**<br>**Cancer deaths=264** | I1: NE<br>I2: NE<br>I3: NE<br>I4: NE<br>I5: NE | I1: NE<br>I2: NE<br>I3: NE<br>I4: NE<br>I5: NE | I1: NE<br>I2: NE<br>I3: NE<br>I4: NE<br>I5: NE |

Values are the difference in the conditional quantile of survival time (years) for cancer mortality per doubling of metal concentration (95% CI) and p value for each imputation (I1–I5). Deaths from other causes were censored. NE=not estimable. Urinary cadmium and arsenic cohorts were restricted to eGFR ≥60 mL/min per 1.73 m². Adjustment as in table S3.

As=arsenic; Cd=cadmium; CQR=censored quantile regression; eGFR=estimated glomerular filtration rate; Hg=mercury; Pb=lead.

**Table S12. Cardiovascular mortality: cause-censored CQR results by imputation**

| Metal | 10th percentile | 25th percentile | 50th percentile |
|---|---|---|---|
| **Blood lead (Pb)**<br>**N=24021**<br>**Cardiovascular deaths=2233** | I1: −0.761 (−1.422, −0.111); p=0.0229<br>I2: −0.734 (−1.480, −0.017); p=0.0494<br>I3: −0.640 (−1.184, −0.134); p=0.0169<br>I4: −0.803 (−1.473, −0.113); p=0.0207<br>I5: −0.709 (−1.273, −0.205); p=0.0093 | I1: −0.764 (−1.476, 0.018); p=0.0449<br>I2: −0.706 (−1.363, −0.037); p=0.0368<br>I3: −0.874 (−1.287, −0.419); p<0.0001<br>I4: −0.765 (−1.516, 0.348); p=0.1074<br>I5: NE | I1: NE<br>I2: NE<br>I3: NE<br>I4: NE<br>I5: NE |
| **Blood mercury (Hg)**<br>**N=18258**<br>**Cardiovascular deaths=1410** | I1: 0.502 (0.268, 0.820); p=0.0004<br>I2: 0.534 (0.131, 0.917); p=0.0078<br>I3: 0.524 (0.078, 1.055); p=0.0355<br>I4: NE<br>I5: 0.550 (0.127, 0.937); p=0.0078 | I1: NE<br>I2: NE<br>I3: NE<br>I4: NE<br>I5: 0.611 (0.159, 1.041); p=0.0066 | I1: NE<br>I2: NE<br>I3: NE<br>I4: NE<br>I5: NE |
| **Urinary cadmium (Cd)**<br>**N=7061**<br>**Cardiovascular deaths=448** | I1: NE<br>I2: −0.557 (−2.385, 1.860); p=0.6067<br>I3: NE<br>I4: NE<br>I5: NE | I1: NE<br>I2: NE<br>I3: NE<br>I4: NE<br>I5: NE | I1: NE<br>I2: NE<br>I3: NE<br>I4: NE<br>I5: NE |
| **Urinary arsenic (As)**<br>**N=5552**<br>**Cardiovascular deaths=283** | I1: NE<br>I2: NE<br>I3: NE<br>I4: NE<br>I5: NE | I1: NE<br>I2: NE<br>I3: NE<br>I4: NE<br>I5: NE | I1: NE<br>I2: NE<br>I3: NE<br>I4: NE<br>I5: NE |

Values are the difference in the conditional quantile of survival time (years) for cardiovascular mortality per doubling of metal concentration (95% CI) and p value for each imputation (I1–I5). Deaths from other causes were censored. NE=not estimable. Urinary cadmium and arsenic cohorts were restricted to eGFR ≥60 mL/min per 1.73 m². Adjustment as in table S3.

As=arsenic; Cd=cadmium; CQR=censored quantile regression; eGFR=estimated glomerular filtration rate; Hg=mercury; Pb=lead.

**Table S13. Cohorts for the sensitivity analyses of the urinary metals**

| Sensitivity | Metal | N / deaths | Excluded: missing eGFR | Excluded: eGFR <60 | Imputation |
|---|---|---|---|---|---|
| **Portnoy** | Cd | 7061 / 1510 | 432 | 973 | Primary imputations |
| **Portnoy** | As | 5552 / 960 | 338 | 779 | Primary imputations |
| **Age ≥55** | Cd | 3982 / 1281 | 265 | 915 | Separate imputation |
| **Age ≥55** | As | 3139 / 837 | 204 | 734 | Separate imputation |
| **Mutual Pb–Cd adjustment** | Pb | 7057 / 1510 | 150 | 971 | Separate imputation |
| **Mutual Pb–Cd adjustment** | Cd | 7057 / 1510 | 150 | 971 | Separate imputation |

Counts are unweighted. All cohorts were restricted to observed eGFR ≥60 mL/min per 1.73 m² before imputation. The age ≥55 years restriction was also applied before imputation. Exclusions are counted relative to the corresponding cohort without the eGFR restriction. The two mutual-adjustment models share the same participants and cadmium sampling weights. eGFR is a covariate only in the cadmium model.

As=arsenic; Cd=cadmium; eGFR=estimated glomerular filtration rate; Pb=lead.

**Table S14. Portnoy estimator**

| Metal | 10th percentile | 25th percentile | 50th percentile |
|---|---|---|---|
| **Blood lead (Pb)**<br>**N=24021; deaths=6885** | I1: −0.646 (−0.859, −0.418); p<0.0001<br>I2: −0.679 (−0.921, −0.435); p<0.0001<br>I3: −0.698 (−0.945, −0.467); p<0.0001<br>I4: −0.699 (−0.949, −0.471); p<0.0001<br>I5: −0.613 (−0.876, −0.352); p<0.0001 | I1: −0.548 (−0.892, −0.172); p=0.0029<br>I2: −0.608 (−0.786, −0.368); p<0.0001<br>I3: −0.621 (−0.868, −0.379); p<0.0001<br>I4: −0.641 (−0.895, −0.428); p<0.0001<br>I5: −0.618 (−0.929, −0.321); p<0.0001 | I1: −0.647 (−0.953, −0.339); p<0.0001<br>I2: −0.625 (−0.802, −0.418); p<0.0001<br>I3: −0.617 (−0.862, −0.338); p<0.0001<br>I4: −0.652 (−0.931, −0.373); p<0.0001<br>I5: −0.630 (−0.966, −0.364); p<0.0001 |
| **Blood mercury (Hg)**<br>**N=18258; deaths=4358** | I1: 0.334 (0.129, 0.529); p=0.0011<br>I2: 0.286 (0.118, 0.435); p=0.0004<br>I3: 0.328 (0.190, 0.461); p<0.0001<br>I4: 0.286 (0.157, 0.422); p<0.0001<br>I5: 0.326 (0.172, 0.491); p<0.0001 | I1: 0.470 (0.326, 0.625); p<0.0001<br>I2: 0.517 (0.322, 0.691); p<0.0001<br>I3: 0.506 (0.320, 0.675); p<0.0001<br>I4: 0.506 (0.317, 0.676); p<0.0001<br>I5: 0.525 (0.266, 0.752); p<0.0001 | I1: 0.578 (0.403, 0.783); p<0.0001<br>I2: 0.480 (0.232, 0.788); p=0.0007<br>I3: 0.520 (0.322, 0.697); p<0.0001<br>I4: 0.591 (0.351, 0.781); p<0.0001<br>I5: 0.535 (0.355, 0.679); p<0.0001 |
| **Urinary cadmium (Cd)**<br>**N=7061; deaths=1510** | I1: −1.752 (−2.381, −1.086); p<0.0001<br>I2: −1.693 (−2.500, −1.074); p<0.0001<br>I3: −1.687 (−2.344, −1.002); p<0.0001<br>I4: −1.715 (−2.223, −1.148); p<0.0001<br>I5: −1.876 (−2.580, −1.278); p<0.0001 | I1: −1.576 (−2.210, −0.972); p<0.0001<br>I2: −1.535 (−2.034, −0.975); p<0.0001<br>I3: −1.498 (−2.053, −0.769); p<0.0001<br>I4: −1.576 (−2.146, −0.927); p<0.0001<br>I5: −1.597 (−2.107, −1.057); p<0.0001 | I1: −1.254 (−1.838, −0.653); p<0.0001<br>I2: −1.466 (−1.988, −0.994); p<0.0001<br>I3: −1.437 (−1.993, −0.808); p<0.0001<br>I4: −1.497 (−2.191, −0.767); p<0.0001<br>I5: −1.351 (−2.150, −0.437); p=0.0020 |
| **Urinary arsenic (As)**<br>**N=5552; deaths=960** | I1: 0.168 (−0.368, 0.722); p=0.5466<br>I2: 0.323 (−0.225, 0.920); p=0.2692<br>I3: 0.047 (−0.404, 0.556); p=0.8476<br>I4: 0.030 (−0.512, 0.611); p=0.9161<br>I5: 0.079 (−0.463, 0.602); p=0.7711 | I1: 0.386 (−0.358, 1.280); p=0.3562<br>I2: 0.436 (−0.404, 1.180); p=0.2812<br>I3: 0.493 (−0.152, 1.258); p=0.1707<br>I4: 0.563 (−0.039, 1.150); p=0.0634<br>I5: 0.424 (−0.378, 1.210); p=0.2956 | I1: 0.416 (−0.275, 1.104); p=0.2377<br>I2: 0.159 (−0.491, 0.947); p=0.6637<br>I3: 0.243 (−0.666, 1.145); p=0.5982<br>I4: 0.155 (−0.980, 1.439); p=0.8013<br>I5: 0.200 (−0.484, 1.098); p=0.6197 |

Values are the difference in the conditional quantile of survival time (years) per doubling of metal concentration (95% CI) and p value for each imputation (I1–I5). 95% CIs were estimated from 400 delete-d jackknife resamples. Urinary cadmium and arsenic cohorts were restricted to eGFR ≥60 mL/min per 1.73 m². Adjustment as in table S3. The Portnoy alternative estimator was applied to the same imputed datasets as the primary analysis using Peng-Huang.

As=arsenic; Cd=cadmium; eGFR=estimated glomerular filtration rate; Hg=mercury; Pb=lead.

**Table S15. Participants aged ≥55 years**

| Metal | 10th percentile | 25th percentile | 50th percentile |
|---|---|---|---|
| **Blood lead (Pb)**<br>**N=14735; deaths=6158** | I1: −0.284 (−0.472, −0.137); p=0.0009<br>I2: −0.315 (−0.510, −0.113); p=0.0019<br>I3: −0.256 (−0.392, −0.089); p=0.0009<br>I4: −0.287 (−0.507, −0.061); p=0.0115<br>I5: −0.273 (−0.552, 0.006); p=0.0548 | I1: −0.342 (−0.638, −0.133); p=0.0079<br>I2: −0.324 (−0.629, −0.020); p=0.0374<br>I3: NE<br>I4: −0.355 (−0.644, −0.084); p=0.0130<br>I5: −0.382 (−0.594, −0.145); p=0.0009 | I1: −0.481 (−0.740, −0.211); p=0.0004<br>I2: −0.446 (−0.726, −0.138); p=0.0030<br>I3: NE<br>I4: NE<br>I5: NE |
| **Blood mercury (Hg)**<br>**N=11187; deaths=3951** | I1: 0.213 (0.088, 0.356); p=0.0018<br>I2: 0.232 (0.055, 0.398); p=0.0081<br>I3: 0.229 (0.087, 0.359); p=0.0010<br>I4: 0.219 (0.115, 0.332); p<0.0001<br>I5: 0.217 (0.047, 0.351); p=0.0051 | I1: 0.455 (0.298, 0.616); p<0.0001<br>I2: 0.426 (0.251, 0.590); p<0.0001<br>I3: 0.385 (0.257, 0.518); p<0.0001<br>I4: 0.441 (0.245, 0.667); p<0.0001<br>I5: 0.423 (0.211, 0.592); p<0.0001 | I1: 0.494 (0.201, 0.783); p=0.0009<br>I2: 0.476 (0.217, 0.671); p<0.0001<br>I3: 0.522 (0.365, 0.695); p<0.0001<br>I4: 0.514 (0.332, 0.694); p<0.0001<br>I5: 0.493 (0.273, 0.664); p<0.0001 |
| **Urinary cadmium (Cd)**<br>**N=3982; deaths=1281** | I1: −1.148 (−1.780, −0.523); p=0.0003<br>I2: −1.253 (−1.963, −0.605); p=0.0003<br>I3: −1.333 (−1.930, −0.741); p<0.0001<br>I4: −1.097 (−1.643, −0.507); p=0.0002<br>I5: −1.166 (−1.733, −0.497); p=0.0002 | I1: −1.433 (−2.068, −0.807); p<0.0001<br>I2: −1.537 (−2.411, −0.570); p=0.0011<br>I3: −1.527 (−2.185, −0.754); p<0.0001<br>I4: −1.323 (−2.126, −0.587); p=0.0007<br>I5: −1.504 (−2.081, −0.873); p<0.0001 | I1: −1.150 (−1.776, −0.541); p=0.0003<br>I2: NE<br>I3: −1.201 (−1.839, −0.611); p=0.0001<br>I4: −1.236 (−1.790, −0.663); p<0.0001<br>I5: −1.160 (−1.831, −0.577); p=0.0003 |
| **Urinary arsenic (As)**<br>**N=3139; deaths=837** | I1: 0.177 (−0.626, 0.829); p=0.6340<br>I2: 0.502 (0.011, 0.995); p=0.0454<br>I3: 0.203 (−0.366, 0.759); p=0.4785<br>I4: 0.288 (−0.210, 0.746); p=0.2376<br>I5: 0.211 (−0.180, 0.683); p=0.3382 | I1: 0.725 (0.104, 1.336); p=0.0211<br>I2: 0.440 (−0.317, 1.009); p=0.1936<br>I3: 0.511 (−0.182, 1.198); p=0.1468<br>I4: 0.553 (−0.210, 1.315); p=0.1551<br>I5: 0.651 (0.044, 1.334); p=0.0478 | I1: 0.308 (−0.394, 1.156); p=0.4355<br>I2: 0.283 (−0.891, 1.457); p=0.6366<br>I3: 0.365 (−0.424, 1.241); p=0.3897<br>I4: 0.319 (−0.347, 0.826); p=0.2859<br>I5: 0.352 (−0.769, 1.603); p=0.5608 |

Values are the difference in the conditional quantile of survival time (years) per doubling of metal concentration (95% CI) and p value for each imputation (I1–I5). NE=not estimable. 95% CIs were estimated from 400 delete-d jackknife resamples. Urinary cadmium and arsenic cohorts were restricted to eGFR ≥60 mL/min per 1.73 m². Adjustment as in table S3.

As=arsenic; Cd=cadmium; eGFR=estimated glomerular filtration rate; Hg=mercury; Pb=lead.

**Table S16. Mutual adjustment for blood lead and urinary cadmium**

| Metal | 10th percentile | 25th percentile | 50th percentile |
|---|---|---|---|
| **Blood lead (Pb)**<br>**N=7057; deaths=1510** | I1: 0.685 (−0.016, 1.456); p=0.0680<br>I2: 0.592 (−0.047, 1.319); p=0.0896<br>I3: 0.590 (−0.105, 1.533); p=0.1580<br>I4: 0.665 (−0.234, 1.625); p=0.1611<br>I5: 0.668 (−0.014, 1.378); p=0.0602 | I1: 0.112 (−0.408, 0.786); p=0.7125<br>I2: 0.254 (−0.587, 0.978); p=0.5244<br>I3: 0.377 (−0.332, 0.898); p=0.2303<br>I4: 0.268 (−0.270, 0.973); p=0.3985<br>I5: 0.129 (−0.945, 0.964); p=0.7916 | I1: 0.314 (−0.292, 0.848); p=0.2801<br>I2: 0.393 (−0.125, 0.859); p=0.1180<br>I3: 0.316 (−0.099, 0.754); p=0.1467<br>I4: 0.153 (−0.413, 0.697); p=0.5879<br>I5: 0.228 (−0.304, 0.786); p=0.4116 |
| **Urinary cadmium (Cd)**<br>**N=7057; deaths=1510** | I1: −1.913 (−2.477, −1.315); p<0.0001<br>I2: −1.810 (−2.566, −1.097); p<0.0001<br>I3: −1.777 (−2.556, −0.804); p<0.0001<br>I4: −1.901 (−2.627, −1.042); p<0.0001<br>I5: −2.035 (−2.794, −1.327); p<0.0001 | I1: −1.583 (−2.072, −0.901); p<0.0001<br>I2: −1.655 (−2.139, −1.115); p<0.0001<br>I3: −1.598 (−2.203, −1.068); p<0.0001<br>I4: −1.637 (−1.927, −1.228); p<0.0001<br>I5: −1.541 (−2.077, −0.840); p<0.0001 | I1: −1.411 (−2.125, −0.525); p=0.0005<br>I2: −1.354 (−2.121, −0.554); p=0.0007<br>I3: −1.298 (−2.021, −0.678); p=0.0002<br>I4: −1.513 (−2.028, −0.990); p<0.0001<br>I5: −1.330 (−2.059, −0.749); p<0.0001 |

Values are the difference in the conditional quantile of survival time (years) per doubling of metal concentration (95% CI) and p value for each imputation (I1–I5). 95% CIs were estimated from 400 delete-d jackknife resamples. Cohorts were restricted to eGFR ≥60 mL/min per 1.73 m². $Log_2$ lead and $log_2$ cadmium entered each model as additive terms (no product term). Both models were adjusted as in table S3. The cadmium model included continuous eGFR.

Cd=cadmium; eGFR=estimated glomerular filtration rate; Pb=lead.

**Table S17. Summary of the sensitivity analyses**

| Sensitivity analysis: metal | 10th percentile | 25th percentile | 50th percentile |
|---|---|---|---|
| **Portnoy: blood lead<br>N=24021; deaths=6885** | −0.699 to −0.613<br>5/5 estimable; 5 CIs exclude 0 | −0.641 to −0.548<br>5/5 estimable; 5 CIs exclude 0 | −0.652 to −0.617<br>5/5 estimable; 5 CIs exclude 0 |
| **Portnoy: blood mercury<br>N=18258; deaths=4358** | 0.286 to 0.334<br>5/5 estimable; 5 CIs exclude 0 | 0.470 to 0.525<br>5/5 estimable; 5 CIs exclude 0 | 0.480 to 0.591<br>5/5 estimable; 5 CIs exclude 0 |
| **Portnoy: urinary cadmium<br>N=7061; deaths=1510** | −1.876 to −1.687<br>5/5 estimable; 5 CIs exclude 0 | −1.597 to −1.498<br>5/5 estimable; 5 CIs exclude 0 | −1.497 to −1.254<br>5/5 estimable; 5 CIs exclude 0 |
| **Portnoy: urinary arsenic<br>N=5552; deaths=960** | 0.030 to 0.323<br>5/5 estimable; 0 CIs exclude 0 | 0.386 to 0.563<br>5/5 estimable; 0 CIs exclude 0 | 0.155 to 0.416<br>5/5 estimable; 0 CIs exclude 0 |
| **Age ≥55 years: blood lead<br>N=14735; deaths=6158** | −0.315 to −0.256<br>5/5 estimable; 4 CIs exclude 0 | −0.382 to −0.324<br>4/5 estimable; 4 CIs exclude 0 | −0.481 to −0.446<br>2/5 estimable; 2 CIs exclude 0 |
| **Age ≥55 years: blood mercury<br>N=11187; deaths=3951** | 0.213 to 0.232<br>5/5 estimable; 5 CIs exclude 0 | 0.385 to 0.455<br>5/5 estimable; 5 CIs exclude 0 | 0.476 to 0.522<br>5/5 estimable; 5 CIs exclude 0 |
| **Age ≥55 years: urinary cadmium<br>N=3982; deaths=1281** | −1.333 to −1.097<br>5/5 estimable; 5 CIs exclude 0 | −1.537 to −1.323<br>5/5 estimable; 5 CIs exclude 0 | −1.236 to −1.150<br>4/5 estimable; 4 CIs exclude 0 |
| **Age ≥55 years: urinary arsenic<br>N=3139; deaths=837** | 0.177 to 0.502<br>5/5 estimable; 1 CI excludes 0 | 0.440 to 0.725<br>5/5 estimable; 2 CIs exclude 0 | 0.283 to 0.365<br>5/5 estimable; 0 CIs exclude 0 |
| **Mutual adjustment: blood lead<br>N=7057; deaths=1510** | 0.590 to 0.685<br>5/5 estimable; 0 CIs exclude 0 | 0.112 to 0.377<br>5/5 estimable; 0 CIs exclude 0 | 0.153 to 0.393<br>5/5 estimable; 0 CIs exclude 0 |
| **Mutual adjustment: urinary cadmium<br>N=7057; deaths=1510** | −2.035 to −1.777<br>5/5 estimable; 5 CIs exclude 0 | −1.655 to −1.541<br>5/5 estimable; 5 CIs exclude 0 | −1.513 to −1.298<br>5/5 estimable; 5 CIs exclude 0 |
| **EPA plus DHA adjustment: blood mercury<br>N=18258; deaths=4358** | 0.272 to 0.334<br>4/5 estimable; 4 CIs exclude 0 | 0.471 to 0.526<br>4/5 estimable; 4 CIs exclude 0 | 0.544 to 0.550<br>3/5 estimable; 3 CIs exclude 0 |

Values are the range of point estimates across imputations (years of conditional survival time per doubling of metal concentration), the number of imputations with an estimate, and the number of 95% CIs excluding zero. Urinary cadmium and arsenic cohorts and the mutual-adjustment cohort were restricted to eGFR ≥60 mL/min per 1.73 m². Imputation-specific estimates are given in tables S14–S16 and S19.

DHA=docosahexaenoic acid; eGFR=estimated glomerular filtration rate; EPA=eicosapentaenoic acid.

**Table S18. Summary of the cancer and cardiovascular mortality results**

| Endpoint: metal | 10th percentile | 25th percentile | 50th percentile |
|---|---|---|---|
| **Cancer: blood lead N=24021; deaths=1503** | −0.947 to −0.726<br>5/5 estimable; 4 CIs exclude 0 | NE (0/5) | NE (0/5) |
| **Cancer: blood mercury N=18258; deaths=987** | 0.493 to 0.580<br>5/5 estimable; 1 CI excludes 0 | NE (0/5) | NE (0/5) |
| **Cancer: urinary cadmium N=7061; deaths=391** | −2.339 to −2.017<br>5/5 estimable; 4 CIs exclude 0 | NE (0/5) | NE (0/5) |
| **Cancer: urinary arsenic N=5552; deaths=264** | NE (0/5) | NE (0/5) | NE (0/5) |
| **Cardiovascular: blood lead N=24021; deaths=2233** | −0.803 to −0.640<br>5/5 estimable; 5 CIs exclude 0 | −0.874 to −0.706<br>4/5 estimable; 2 CIs exclude 0 | NE (0/5) |
| **Cardiovascular: blood mercury N=18258; deaths=1410** | 0.502 to 0.550<br>4/5 estimable; 4 CIs exclude 0 | 0.611<br>1/5 estimable; 1 CI excludes 0 | NE (0/5) |
| **Cardiovascular: urinary cadmium N=7061; deaths=448** | −0.557<br>1/5 estimable; 0 CIs exclude 0 | NE (0/5) | NE (0/5) |
| **Cardiovascular: urinary arsenic N=5552; deaths=283** | NE (0/5) | NE (0/5) | NE (0/5) |

Values are the range of point estimates across imputations (years of conditional survival time per doubling of metal concentration, with deaths from other causes censored), the number of imputations with an estimate, and the number of 95% CIs excluding zero. NE=not estimable in any imputation. Urinary cadmium and arsenic cohorts were restricted to eGFR ≥60 mL/min per 1.73 m². Imputation-specific results are given in tables S11 and S12.

eGFR=estimated glomerular filtration rate.

**Table S19. Blood mercury: adjustment for dietary EPA plus DHA intake in place of fish intake**

| Metal | 10th percentile | 25th percentile | 50th percentile |
|---|---|---|---|
| **Blood mercury (Hg)**<br>**N=18258; deaths=4358** | I1: 0.322 (0.183, 0.475); p<0.0001<br>I2: 0.334 (0.199, 0.472); p<0.0001<br>I3: 0.272 (0.131, 0.456); p=0.0010<br>I4: 0.308 (0.117, 0.497); p=0.0015<br>I5: NE | I1: 0.501 (0.312, 0.714); p<0.0001<br>I2: 0.481 (0.222, 0.736); p=0.0002<br>I3: 0.526 (0.310, 0.783); p<0.0001<br>I4: 0.471 (0.208, 0.730); p=0.0004<br>I5: NE | I1: 0.550 (0.321, 0.762); p<0.0001<br>I2: NE<br>I3: 0.544 (0.362, 0.771); p<0.0001<br>I4: 0.547 (0.385, 0.690); p<0.0001<br>I5: NE |

Values are the difference in the conditional quantile of survival time (years) per doubling of metal concentration (95% CI) and p value for each imputation (I1–I5). NE=not estimable. 95% CIs were estimated from 400 delete-d jackknife resamples. Adjustment as in table S3, with dietary EPA plus DHA intake in place of fish intake.

DHA=docosahexaenoic acid; EPA=eicosapentaenoic acid; Hg=mercury.

### Additional methods: exposure measurement and urinary dilution

These details describe the continuous-NHANES primary analyses. The outcome cohorts remain 1999–2014 for lead and cadmium and 2003–2014 for mercury and arsenic. The later creatinine-reference cycles described below do not extend the outcome cohorts.

Blood lead (LBXBPB, µg/dL) was measured by graphite-furnace atomic absorption spectrometry in 1999–2002 and by ICP-MS from 2003. Total blood mercury (LBXTHG, µg/L) in the 2003–2014 cohort was measured by ICP-MS. Urinary cadmium (URXUCD, µg/L) used ICP-MS methods. Urinary arsenic species used high-performance liquid chromatography coupled to ICP-MS with a dynamic reaction cell. NHANES released laboratory quality-control procedures and cycle-specific documentation. Selected source documents are linked below. We retained released concentrations and below-detection substitutions. Survey-cycle indicators were included in outcome models. These do not eliminate all potential assay differences between cycles.

The arsenic exposure was URXUAS3 + URXUAS5 + URXUMMA + URXUDMA (arsenous acid, arsenic acid, monomethylarsonic acid, and dimethylarsinic acid). All four species had to be observed. A missing species made the sum missing. Arsenobetaine was excluded. Each urinary concentration was standardised for dilution before $\log_2$ transformation. Blood concentrations were $\log_2$-transformed without urinary-creatinine standardisation.

### Creatinine standardisation

An unweighted ordinary least-squares model fitted natural-log urinary creatinine (URXUCR, mg/dL) as a function of baseline age, age$^2$, sex, race and ethnicity, BMI, and categorical survey cycle. It used 28,555 complete records from the retained 1999–2018 reference dataset. Predicted creatinine was the exponentiated linear predictor, without a smearing factor. Standardised metal concentration = measured metal concentration × predicted creatinine / measured creatinine. The reference coefficients were fixed, rather than refitted for each metal, outcome, imputation, or resample. Baseline urinary creatinine in 2001–2002 came from L16_B. Repeat examinations were not averaged with baseline.

A finite positive standardised concentration was required for inclusion. Missing prediction inputs, including BMI, could therefore exclude a participant even when the urinary metal was measured. Urinary creatinine was not entered again in the outcome regression. This dilution correction is distinct from the eGFR eligibility restriction and adjustment. A model using the transformed urinary exposure is not an analysis of the raw urine concentration.

### Source documentation and original-survey support

CDC/NCHS laboratory documentation: blood metals 1999–2000; 2001–2002; 2003–2004; 2013–2014; urinary metals 2013–2014; arsenic species 2003–2004; 2013–2014

NCHS within CDC sponsored and conducted the original NHANES surveys, including survey design, data collection, and public release. The present secondary analysis received no funding. CDC/NCHS had no role in its design, analysis, interpretation, writing, or submission. Available eligible participants determined study size. No formal sample-size calculation was performed.

Survey provenance: CDC: About NHANES; NIAAA data directory: sponsoring agency for NHANES III and continuous NHANES (accessed Nov 11, 2026).

### Table S20. Operational definitions of the primary adjustment variables

| Variable | Source / analysis field | Definition and model coding |
|---|---|---|
| Age; sex | Demographics: RIDAGEYR; RIAGENDR | Age in years, entered as linear and quadratic terms, retaining released top-coding. Sex categories: male (1) and female (2). |
| Race and ethnicity | Demographics: RIDRETH1 | Mexican American (1), other Hispanic (2), non-Hispanic White (3), non-Hispanic Black (4), other (5). |
| Education | Demographics: DMDEDUC2 | Less than 9th grade; 9th–11th grade, including 12th grade without diploma; high-school graduate/GED; some college/associate degree; college graduate or above (codes 1–5). Codes 7/9 were missing. |
| Marital status | Demographics: DMDMARTL | Married/cohabiting (1/6); widowed/divorced/separated (2/3/4); never married (5). Codes 77/99 were missing. |
| Income-to-poverty ratio | Demographics: INDFMPIR | Continuous ratio of family income to poverty threshold. Released top-coding was retained. |
| BMI | MEC examination: BMXBMI | Continuous baseline BMI, kg/m². The baseline 2001–2002 measurement was used, not an average including the repeat examination. |
| Smoking | Inherited smoking questionnaire fields: pack_years | Continuous pack-years. Observed values equal cigarettes/day ÷ 20 × years smoked. Original harmonisation and missingness rules were incompletely recoverable. In 290 reference records with finite arithmetic inputs, pack-years remained missing. |
| Alcohol use | Inherited alcohol questionnaire fields: alq_5cat | Lifetime abstainer, former drinker, light, moderate, heavy. Sex-specific intensity thresholds and missingness gates are specified on p 28. No additional binge-drinking criterion entered this category. |
| Physical activity | Inherited activity questionnaire field: pa_mets_wkly_all | Four unweighted, cycle-specific rank groups of the stored weekly MET measure (p 28). Original cross-cycle MET derivation was unavailable. Ties can cross group boundaries. |
| Survey cycle | Demographics: SDDSRVYR | Categorical indicators for cycles 1–8 (1999–2014) in lead/cadmium and 3–8 (2003–2014) in mercury/arsenic models. |
| Total cholesterol | Laboratory: LBXTC | Continuous baseline serum total cholesterol, mg/dL. The 2001–2002 repeat-examination values were not substituted or averaged. |
| Kidney function | Baseline calibrated serum creatinine: LBXSCR_cal | 2021 race-free CKD-EPI eGFR, mL/min per 1.73 m². Required observed eGFR ≥60 before imputation for urinary-metal models and adjusted continuously thereafter. Baseline 2001–2002 creatinine was native LBDSCR from L40_B without additional calibration. |
| Fish intake | Dietary screener/recall: fish_servings_wk | Continuous servings/week, mercury model only. The priority hierarchy and cohort-specific counts are on p 29. |

Categorical variables were unordered factors, with the first category listed as reference. Physical activity used Q1, and survey cycle used the earliest included cycle. Other covariates remained continuous. Categories in descriptive tables did not replace continuous model terms. The DAG informed inclusion. Inherited behavioural categories were retained to represent intake/activity differences without a linear category-score assumption. Their historical choice of thresholds was not independently recoverable.

eGFR = 142 × min(Scr/k, 1)^a × max(Scr/k, 1)^−1.2 × 0.9938^age × F. For females, k=0.7, a=−0.241, F=1.012. For males, k=0.9, a=−0.302, F=1. Scr is serum creatinine in mg/dL. The equation is cited in the main manuscript (reference 19). Missing observed eGFR was an eligibility exclusion, not an imputed value for the primary urinary cohorts.

### Table S21. Alcohol thresholds and observed physical-activity group ranges

Alcohol categories reproduced the stored coding as follows: missing ever-drinker status gave a missing category. A value of ever_drinker=0 indicated lifetime abstention. Among ever drinkers, missing annual drinking days gave missing alcohol status, and zero annual drinking days indicated former drinking. For current drinkers with an observed daily average, male light/moderate/heavy thresholds were <2, ≥2 to <3, and ≥3 drinks/day. Female thresholds were <1, ≥1 to <2, and ≥2 drinks/day. A missing daily average remained missing.

Among jointly observed inputs, annual drinking days equalled ALQ120Q multiplied by 52, 12, or 1 for weekly, monthly, or yearly frequency (ALQ120U=1, 2, or 3). Average drinks/day equalled ALQ130 × annual drinking days / 365. These verified mappings do not recover all upstream questionnaire cleaning or the definition of the inherited ever_drinker field. Original missing values were retained.

Activity groups were defined in the retained 24,339-person through-2014 reference dataset, before removal of one thallium-only participant or selection of a metal cohort. Finite weekly MET values were sorted within each cycle, with ties ordered by archive row position, then split into four consecutive, nearly equal-sized groups (first groups received any remainder). This reproduces all 23,845 observed assignments and their missingness. These are rank groups, not disjoint exposure intervals: zero values cross Q1/Q2 boundaries, and also Q2/Q3 in 1999–2000. They were not re-estimated within each metal cohort.

| Survey cycle | Q1: min–max (n) | Q2: min–max (n) | Q3: min–max (n) | Q4: min–max (n) |
|---|---|---|---|---|
| 1999–2000 | 0–0 (n=663) | 0–0 (n=663) | 0–735 (n=663) | 735–33600 (n=663) |
| 2001–2002 | 0–0 (n=737) | 0–122.5 (n=736) | 122.5–981.75 (n=736) | 982.333–21294 (n=736) |
| 2003–2004 | 0–0 (n=712) | 0–106.167 (n=712) | 106.167–816.667 (n=711) | 817.833–35732.7 (n=711) |
| 2005–2006 | 0–0 (n=681) | 0–147 (n=681) | 147–910 (n=680) | 910–24990 (n=680) |
| 2007–2008 | 0–0 (n=925) | 0–600 (n=925) | 600–3120 (n=924) | 3120–55440 (n=924) |
| 2009–2010 | 0–0 (n=970) | 0–720 (n=969) | 720–2880 (n=969) | 2880–55440 (n=969) |
| 2011–2012 | 0–0 (n=824) | 0–720 (n=824) | 720–2760 (n=824) | 2800–51840 (n=824) |
| 2013–2014 | 0–0 (n=453) | 0–720 (n=452) | 720–2560 (n=452) | 2580–50400 (n=452) |

Ranges are in the units of the inherited weekly MET measure (pa_mets_wkly_all). They report the analysed values without asserting that questionnaire-derived activity was fully comparable across cycles. Counts are unweighted, non-missing reference-sample counts, not metal-specific counts. Group membership cannot be reproduced from a numeric boundary alone because ties were split. MET=metabolic equivalent of task.

The grouping algorithm and alcohol cut-offs were verified against the saved data. Incomplete recovery of their original raw-questionnaire construction remains a measurement-reporting limitation. No new thresholds or MET values were substituted in the analyses.

### Table S22. Fish-intake sources in the 2003–2014 mercury cohort

| Source before imputation | Participants | Unweighted percentage of 18,258 |
|---|---|---|
| Tier 1: 30-day species screener | 14,757 | 80.82% |
| Tier 2: reliable 24-h recall fallback | 2,505 | 13.72% |
| Missing before multiple imputation | 996 | 5.46% |
| Total | 18,258 | 100.00% |

Tier 1 comprises 11,298 species-screener records (including supported zero intake of the target species) and 3,459 explicit screener-zero records. Counts for tuna, mackerel, salmon, sardines, and trout in the preceding 30 days were summed and divided by 4.345 to obtain servings/week. A valid observed screener value, including zero, always took priority.

Tier 2 was used only when Tier 1 was unavailable. It comprises 2,326 two-day and 179 one-day reliable dietary recalls. The stored mean grams/day of dark fish was divided by 85 and multiplied by 7 to obtain weekly servings. Residual missing intake was multiply imputed in the mercury cohort. The original recall food-code list and all upstream exclusions were not recoverable from the archived builder. The supplied values and missingness were retained.

These counts use the exact 18,258 participants in the primary mercury model, not the earlier 1999–2018 cohort. Percentages here are unweighted. Table S2 reports the same 996 missing participants as 4.9% after survey weighting. In the dietary sensitivity analysis, EPA plus DHA was (DRXTP205 + DRXTP226) × 1000 mg/day. The inherited recall-day selection was not fully documented.

### Reporting of adjusted estimates

The study objective concerns survival-time quantiles conditional on the specified covariates. Accordingly, only confounder-adjusted CQR estimates are presented (main table 2 and table S3). Unadjusted estimates are omitted because the report focuses on this conditional estimand. This choice prevents direct comparison of crude and adjusted associations. Confounder adjustment alone does not establish causality. No unadjusted estimates are implied by the reported results.

### Bias and uncertainty

Single measurements and imperfect behavioural adjustment can produce material exposure error and residual confounding. Attenuation is plausible for variable biomarkers, but the direction is not guaranteed in multivariable quantile models. Renal restriction/adjustment could remove mediated associations. Preclinical disease and residual smoking confounding could exaggerate inverse cadmium associations. Dietary confounding could account for a positive mercury association. Selection on exposure availability and eGFR could act in either direction. The data do not quantify these bias magnitudes or rule out changes in direction. Sensitivity analyses assess robustness to specified model changes, not the size of unmeasured bias. Confidence intervals do not include systematic bias, and the primary intervals also omit between-imputation uncertainty and complex-survey variance estimation.
